# Enhanced NF-κB signaling in type-2 dendritic cells at baseline predicts non-response to adalimumab in psoriasis

**DOI:** 10.1101/2020.11.09.20228502

**Authors:** Rosa Andres-Ejarque, Hira Bahadur Ale, Katarzyna Grys, Isabella Tosi, Shane Solanky, Chrysanthi Ainali, Zeynep Catak, Hemawtee Sreeneebus, Jake Saklatvala, Nick Dand, Emanuele de Rinaldis, Anna Chapman, Frank O. Nestle, Michael R. Barnes, Richard B. Warren, Nick J. Reynolds, Christopher E.M. Griffiths, Jonathan N. Barker, Catherine H. Smith, Paola Di Meglio, on behalf of the PSORT Consortium

**Author notes:** **Address correspondence** to Paola Di Meglio, St. Johns Institute of Dermatology, Guy’s Hospital, Tower Wing, Great Maze Street, London SE1 9RT. EDR and FON’s present affiliation: Sanofi, Cambridge, Mass, USA. **Authorship note:** RBW, NJR, CEMG, JNB, CHS contributed equally to this work. The members of the PSORT consortium (excluding individually named authors of this work) are Richard Emsley, Andrea Evans, Katherine Payne, and Deborah Stocken. EdR and FON are currently employees of Sanofi. MRB has received honoraria and/or research grants from Janssen, Servier and Lilly. RBW has received honoraria and/or research grants from AbbVie, Almirall, Amgen, Boehringer Ingelheim, Celgene, Janssen, Leo, Lilly, Novartis, Pfizer, Sanofi, Xenoport, and UCB. NJR has received research grants from GSK-Stiefel and Novartis; and other income to Newcastle University from Almirall, Amgen, Janssen, Novartis, Sanofi Genzyme Regeneron and UCB Pharma Ltd for lectures/attendance at advisory boards. CEMG reports grants and/or personal fees from AbbVie, Almirall, Amgen, BMS, Celgene, Galderma, Janssen, Leo Pharma, Lilly, Novartis, Sandoz and UCB Pharma. JNB has received honoraria and/or research grants from AbbVie, Almirall, Amgen, Boehringer-Ingelheim, Bristol Myers Squibb, Celgene, Janssen, Leo, Lilly, Novartis, Samsung, Sun Pharma. CHS has received departmental research funding from AbbVie, GSK, Pfizer, Novartis, Regeneron, and Roche. PDM reports grants and/or personal fees from Janssen, Novartis, and UCB Pharma. All the other authors do not report any competing interest.

## Abstract

Biological therapies have transformed the management of psoriasis but clinical outcome is variable leaving an unmet clinical need for predictive biomarkers of response. To identify the immune determinants of response to the anti-TNF drug adalimumab and evaluate their predictive value, we performed in-depth immunomonitoring of blood immune cells of 67 psoriasis patients, before and during therapy. We assessed proximal TNF signaling events, by measuring NF-κB nuclear translocation and phosphorylation, and downstream effects, such as cell phenotype and function. Enhanced NF-κBp65 phosphorylation, induced by TNF and LPS in type-2 dendritic cells (DC) before therapy, significantly correlated with lack of clinical response after 12 weeks’ treatment. The heightened NF-κB activation was mechanistically linked to increased DC maturation *in vitro* and frequency of IL-17^+^T cells in the blood of non-responders before therapy. Moreover, lesional skin of non-responders contained more activated dermal DC and increased numbers of IL-17^+^T cells. sFinally, we identified and clinically validated LPS-induced NF-κBp65 phosphorylation before therapy as a predictive biomarker of non-response to adalimumab, with 100% sensitivity and 90.1% specificity in an independent cohort. Our study uncovers key molecular and cellular players underpinning adalimumab mechanisms of action in psoriasis and proposes a blood biomarker for predicting clinical outcome.

## Introduction

Psoriasis is a common, chronic, immune-mediated, inflammatory skin disease, affecting over 100 million people worldwide and is recognized as serious noncommunicable disease by the World Health Organization for the significant negative impact it has on people’s lives ^1^.

Painful, disfiguring and disabling skin lesions result from the combination of genetic susceptibility, environmental triggers and dysregulated immune responses involving dendritic cells (DC), IL-17-producing T (T17) cells, keratinocytes and pro-inflammatory cytokines ^2,3^. Over the last two decades, significant advances in understanding the pathogenic mechanisms underpinning psoriasis, have led to the adoption of biological treatments (“biologics”) targeting the key cytokines TNF, IL-23 and IL-17A, which have transformed disease management. Nevertheless, response to biologics is often heterogeneous, with lack or loss of response a persistent issue in up to 30% of patients, in addition to potential side-effect, such as infections and allergic reactions ^4^. Variability of response and treatment failure results in treatment switching, which in turn increases both patients’ dissatisfaction and costs for healthcare providers. Thus, psoriasis could greatly profit from the implementation of stratified medicine approaches to benefit patients and reduce costs ^5^. The choice of which biologic to prescribe is currently based on clinical factors, for example anti-TNF are preferentially prescribed to patients with concomitant psoriatic arthritis^6^. The PSORT consortium, a unique partnership involving clinicians, translational researchers, industry and patient groups, aims to identify determinants of response to biologic therapies, and ultimately deliver a patient stratifier to guide psoriasis management ^7^. The analysis of demographic, social, clinical, pharmacological and genomic data has identified a number of factors, such as ethnicity, weight, smoking ^8^ serum drug lvels ^9^ and HLA-Cw0602 status ^10^, which are associated with outcome to biologics. However, the small effect size suggest that these factors alone are insufficient to inform optimal treatment selection. Thus, there is an unmet clinical need to identify biomarkers predictive of response to biologics. In particular, biomarkers involved in disease pathogenesis and/or drug mechanism of action (i.e. mechanistic biomarkers) are considered more informative, robust and actionable than biomarkers simply reflecting the clinical outcome ^11^.

Anti-TNF were the first class of biologics approved for psoriasis, in line with the pleiotropic role of TNF in inflammation. Activation of the transcription factor nuclear factor-κB (NF-κB) is a key event downstream TNF signaling, inducing the expression of inflammatory and anti-apoptotic genes, which influence critical cellular behaviors such as activation, maturation, migration, proliferation and survival ^12^. Adalimumab is a recombinant, fully humanized, IgG antibody with high affinity and specificity for both soluble and membrane-bound TNF. In clinical trials, 71% of patients with moderate-to-severe psoriasis achieved a 75% reduction in the standard objective disease severity score used in psoriasis - Psoriasis Area and Severity Index (PASI; PASI75 response) after 16 weeks of adalimumab treatment, with 45% achieving 90% reduction (PASI90 response) ^13^. Whilst newer biologics have been developed and licensed, adalimumab remains a first line intervention in psoriasis, given its effectiveness and well-established safety profile and – with the advent of biosimilars - significantly reduced costs. Nevertheless, the cellular and molecular mechanisms underpinning its clinical efficacy are ill understood. In a small early study, adalimumab induced normalization of keratinocyte differentiation and decreased the number of T cells and DC in psoriasis skin lesions ^14^, in keeping with the down-modulation of myeloid and Th17-response genes observed in patients treated with etanercept ^15,16^. Little is known about the effect of adalimumab on circulating blood immune cells in psoriasis. In particular, the effect of TNF-neutralization on proximal signaling events, such as NF-κB activation, has not been investigated thus far. Here, as a fundamental step to identify mechanistic immune determinants of response to adalimumab and evaluate their value as predictive biomarkers, we sought to elucidate the blood cellular target(s) and molecular effect of adalimumab in psoriasis. Thus, we exploited the unprecedented clinical psoriasis bioresource amassed by the PSORT Consortium ^7^ and performed in-depth functional and phenotypic immunomonitoring of blood immune cells of 67 psoriasis patients, before and during biologic therapy, and 20 healthy controls. Next, we validated our findings with mechanistic *in vitro* experiments and skin *in situ* imaging studies. Finally, we compared the predictive value of the most promising immune traits identified and validated the best mechanistic immune biomarker predictive of response in an independent patient cohort.

## Results

### Adalimumab therapy blocks TNF-induced NF-κB translocation in lymphoid cells and, to a lesser extent, in dendritic cells

To obtain mechanistic insights into the cellular target and effect of adalimumab in blood we monitored NF-κB activation in 16 patients undergoing adalimumab treatment (PSORT adalimumab discovery cohort, **Supplemental Figure 1**), measuring NF-κB nuclear translocation in 7 major immune cell populations by imaging flow cytometry. Fresh whole blood, obtained at baseline before starting therapy (week 0, w0) and at week 1 (w1), 4 (w4) and 12 (w12) after the initiation of the treatment, was stimulated with either TNF or LPS or left unstimulated, and NF-κB nuclear translocation was quantified as Rd score in neutrophils, monocytes, DC, natural killer (NK), NKT, CD4+ and CD8+ T cells (**Figure 1a** and **Supplemental Figure 2**). Psoriasis patients receiving the anti-12/23p40 mAb ustekinumab (n=24) and matched healthy volunteers (n=11) were used as control groups (**Supplemental Figure 1**). No difference in NF-κB constitutive nuclear localization, that is in absence of stimulation, was observed at any time point in any cell type (**Supplemental Figure 3a**). Following stimulation, TNF-induced NF-κB translocation in both lymphoid and myeloid cells, while, as expected, LPS only activated neutrophils, monocytes and DC in blood collected at w0 **(Figure 1b,c**). A similar activation profile was observed in whole blood of healthy volunteers (**Supplemental Figure 3b**). NF-κB activation induced by TNF in lymphoid cells was strongly inhibited in the blood of patients receiving adalimumab at w1, w4 and w12 of treatment (**Figure 1b,c**), but not in those receiving ustekinumab (**Supplemental Figure 3c**), in line with the molecular pathway targeted by each drug. Adalimumab exerted the strongest inhibitory effect in lymphoid cells, i.e. CD4^+^T, CD8^+^T, NK and NKT cells, where NF-κB translocation was significantly inhibited by 70-96% (**Figure 1b,c** and **Supplemental Table 1**). In contrast, adalimumab therapy resulted in a weaker inhibition in myeloid cells, with only partial inhibition in DC (w1, 50%, FDR = 0.0061; w4, 50%, FDR = 0.0066; w12, 70 %, FDR = 0.0086), and no effect in monocytes and neutrophils (**Figure 1b,c**, and **Supplemental Table 1**). To confirm this inhibition pattern, we stimulated whole blood obtained from healthy volunteers (n=3) with increasing concentration of TNF, in presence or absence of adalimumab added to the culture. In keeping with the results obtained in patients undergoing adalimumab therapy, TNF-induced NF-κB nuclear translocation was significantly inhibited by up to 98% in T, NK and NKT cells, but only partially in DC (70.48%, FDR = 0.084) with no effect on neutrophils and monocytes (**Supplemental Figure 3d**). As expected, neither adalimumab nor ustekinumab therapy had any effect on LPS-induced NF-κB translocation in psoriasis patients (**Figure 1b,c**; **Supplemental Figure 3c** and **Supplemental Table 1**). Thus, we concluded that the free adalimumab present in the blood stream of patients undergoing therapy completely blocks NF-κB signalling in T, NK and NKT cells but only partially in DC, raising the possibility that the incomplete inhibition observed in DC may affect clinical outcome.

**Figure 1.**
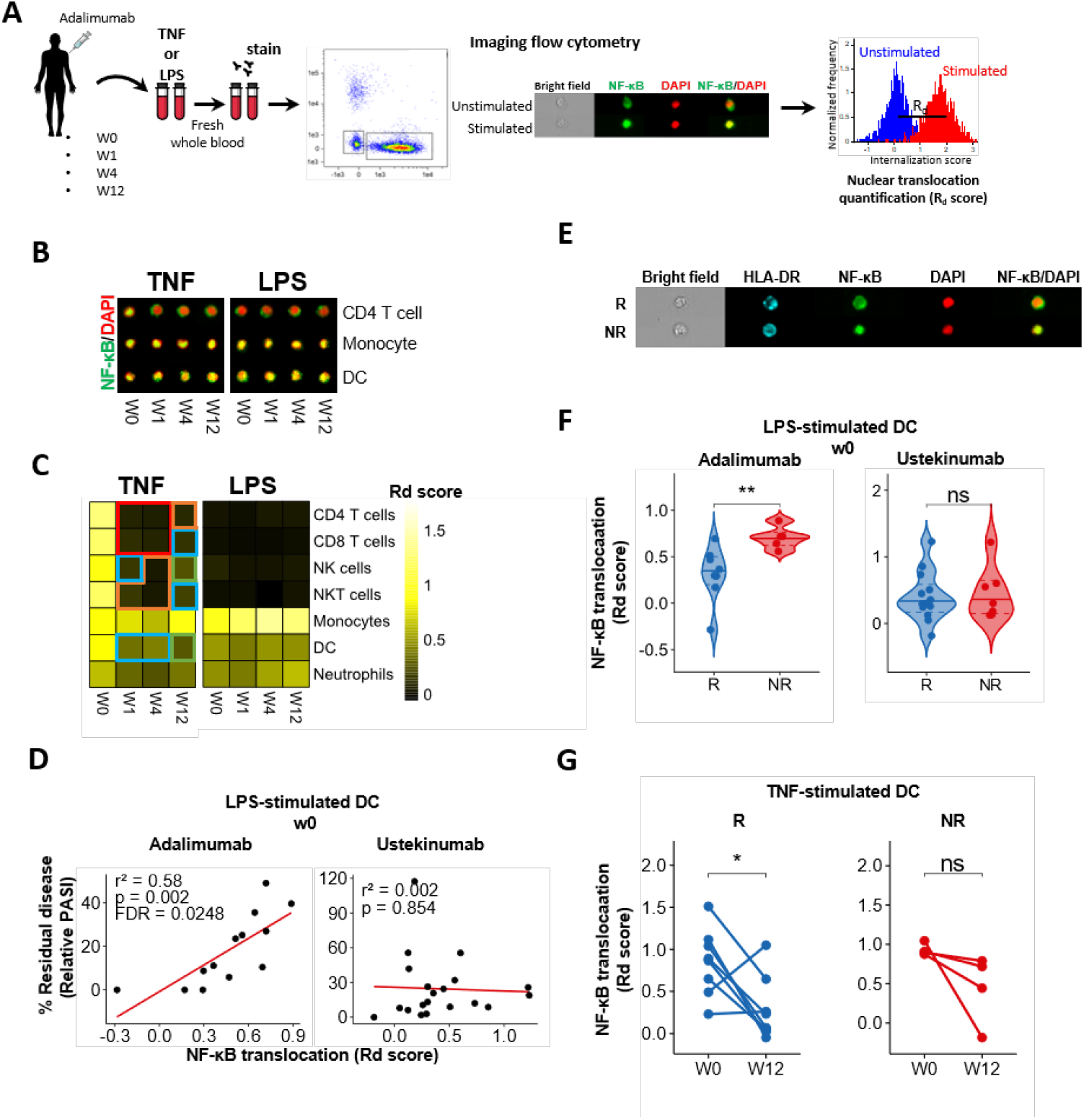
LPS-induced NF-κB translocation in dendritic cells at baseline correlates with lack of response to adalimumab at week 12. **(A)** Whole blood from psoriasis patients collected at baseline (week0, w0) and at w1, w4 and w12 after initiation of adalimumab therapy was stimulated with TNF, LPS or left unstimulated, stained with fluorescent antibodies and NF-κB nuclear translocation was measured in distinct cell subsets, as specified by imaging flow cytometry. **(B)** Representative fluorescent images of selected cell subsets stained for NF-κB (green) and nucleus (DAPI, red); their colocalization is shown in yellow. **(C)** Heatmap showing NF-κB nuclear translocation measured as Rd score in immune cell populations. **(D)** Correlation analysis between NF-κB translocation (measured as Rd score) at w0 and clinical response expressed as percentage of residual disease at w12 (measured as relative PASI, i.e. PASI at w12/ PASI at w0 × 100) in LPS-stimulated dendritic cells (DCs) in PSORT adalimumab (n=13) and ustekinumab cohort (n=24). Each dot represents one patient. **(E)** Representative fluorescent image of LPS-stimulated DCs at w0 from PASI75 adalimumab responder (R) and non-responder (NR). **(F)** Violin plot graphs of NF-κB translocation in LPS-stimulated DCs at w0 for PASI75 adalimumab and ustekinumab R (blue, n_adalimumab_=8; n_ustekinumab_=13) and NR (red, n_adalimumab_=5; n_ustekinumab_=7). (**G**) NF-κB translocation in TNF-stimulated DCs at w0 and w12 in adalimumab PASI75 R (n=8) and NR (n=4). Red frame: FDR < 0.0001, orange frame: FDR < 0.001, blue frame: FDR < 0.01, green frame: FDR < 0.05 compared to week (w) 0, test. ** *p* < 0.01, * *p* < 0.05, Wilcoxon followed by FRD (C), Wilcoxon (G) or Mann-Whitney U test (F).

### NF-κB nuclear translocation in dendritic cells at baseline specifically correlates with lack of clinical response to adalimumab at week 12

Next, we investigated whether NF-κB nuclear translocation, induced by TNF or LPS, correlated with clinical response to adalimumab. Response was assessed applying either a continous model, where the outome is % residual disease at week 12 measured by relative PASI (i.e. PASI at week 12 divided by PASI at w0) x 100), or a binary model, using gold standards PASI75 and PASI90. No correlation between TNF-induced NF-κB nuclear translocation and % residual disease was detected in any cell type at any time point (**Supplemental Table 2**), despite the inhibitory effect of adalimumab in lymphoid cells **(Figure 1c)**. Thus, the complete inhibition of NF-κB signalling in T, NK and NKT cells does not underpin clinical response to adalimumab. Notably, however, we found a statistically significant correlation (r^2^=0.58, p=0.002, FDR < 0.05) between LPS-induced NF-κB translocation at w0 in DCs and % residual disease, with increased NF-κB activation in patients with higher residual disease at w12 (**Figure 1d** and **Supplemental Table 2**). Importantly, this correlation was not observed in ustekinumab-treated patients (**Figure 1d**), indicating that NF-κB activation in DCs is a specific marker of clinical response to adalimumab, and implicates DC NF-κB activation in the cellular and molecular mechanisms underlying adalimumab response. Moreover, LPS-induced NF-κB translocation at w0 was significantly (p < 0.01) lower in DCs of patients achieving PASI75 and PASI90 response (i.e responders, R) than in non-responders (NR) (**Figure 1e, f** and **Supplemental Figure 3e**), while no difference was observed in the ustekinumab cohort (**Figure 1f**). Finally, TNF-induced NF-kB activation in DC at w12 was significantly reduced by adalimumab in R but not in NR (**Figure 1g)**. Thus, NF-κB activation in DC prior to therapy differs in adalimumab R and NR and DC in NR subjects may be more refractory to the inhibitory effect of adalimumab therapy.

### Increased NF-κB phosphorylation in type 2 conventional dendritic cells (cDC2) at baseline correlates with lack of clinical response to adalimumab at week 12

The NF-κB activation cascade includes phosphorylation of p65 NF-kB subunit, such as at Ser529, which regulate NF-κB function ^17^. Thus, we developed a 13-colours phospho-flow cytometry panel (**Supplemental Figure 4**) to study NF-κB phosphorylation in cryopreserved PBMCs of patietns receiving adalimumab. PBMCs were stimulated with LPS or TNF and phopho-p65 was measured as log_10_ fold change (FC) Median Fluorescence Intensity (MFI) in 11 cell subsets, including plasmacytoid dendritic cells, CD141+ type 1 conventional DC (cDC1), CD1c+ type 2 conventional DC (cDC2) and double negative DC (dnDC) (**Figure 2a**). In keeping with the assay being performed in isolated PBMCS rather than in whole blood, and thus in absence of free adalimumab in the blood, we did not detect any inhibitory effect of adalimumab on TNF-induced NF-κB phosphorylation during therapy, in neither lymphoid or myeloid cells **(Supplemental Figure 5** and **Supplemental Table 3)**. Next, we evaluated NF-κB-p65 phosphorylation in PBMCs of the PSORT adalimumab discovery cohort (n=16) previously assayed for NF-κB nuclear translocation. In keeping with the correlation between NF-κB nuclear translocation and residual disease observed in DCs (**Figure 1d**), we detected a statistically significant correlation (r^2^=0.714, p=3×10^−4^, FDR<0.01) between LPS-induced NF-κB phosphorylation in cDC2 at baseline and residual disease at w12 (**Figure 2b and Supplemental Table 4**), thus analytically validating our previous finding using a different, but biologically related, analytical read-out. Moreover, LPS-induced NF-κB phosphorylation at w0 was significantly lower in cDC2 of PASI75 R patients (p<0.05; **Figure 2c**), with a similar downward trend also observed in patients reaching PASI90 (**Supplemental Figure 6a**). NF-κB phosphorylation in cDC2 significantly correlated with NF-κB nuclear translocation observed in total DC (r^2^=0.586, p=0.099, **Figure 2d**), further validating cDC2 as the main DC subset implicated in clinical response to adalimumab.

**Figure 2.**
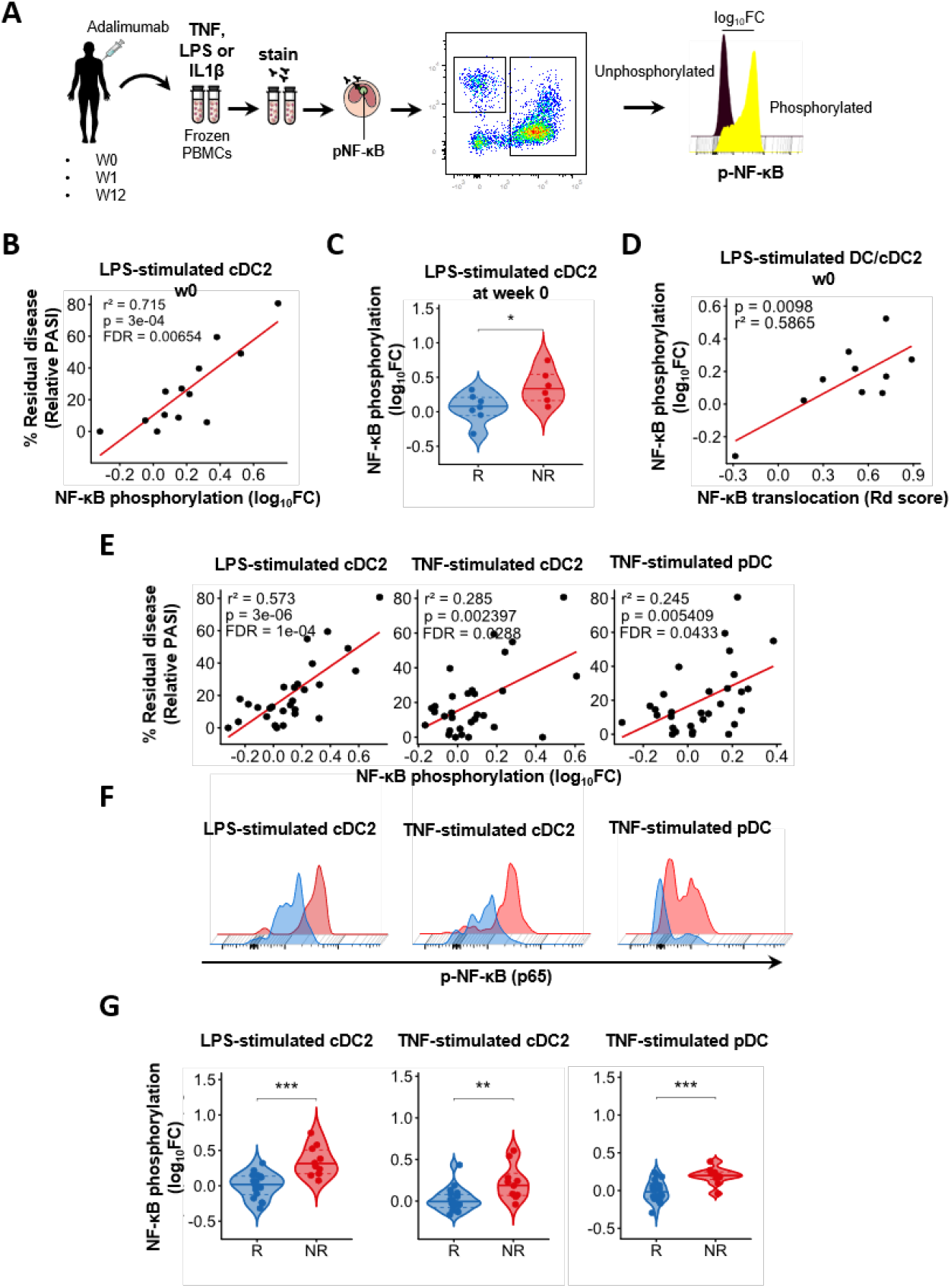
LPS-induced NF-κB phosphorylation in cDC2 at baseline correlates with lack of response to adalimumab. **(A)** PBMC from psoriasis patients obtained at baseline (week 0, w0) and at w1 and w12 after starting adalimumab therapywere stimulated with either TNF or LPS and NF-κB p65 phosphorylation was measured by ph ospho flow cytometry. **(B)** Correlation analysis between LPS-induced NF-κB p65 phosphorylation (log_10_FC) in conventional Type 2 DC (cDC2) at w0 and clinical response, expressed as residual disease at w12 and measured as relative PASI in the discovery cohort (n =13). Each dot represents one patient. **(C)** Violin plot graphs of LPS-induced NF-κB p65 phosphorylation in cDC2 at w0 in PASI75 adalimumab responders PASI (R, blue, n=7) and non-responders (NR, red, n=6). **(D)** Correlation between LPS-induced NF-κB nuclear translocation in DC and LPS-induced NF-κB p65 phosphorylation in cDC2 at w0. **(E)** Correlation analyses between NF-κB p65 phosphorylation induced by different stimuli in cDC2 and plasmacytoid Dc (pDC) and residual disease in the PSORT adalimumab combined cohort (*n* = 25-30). **(F)** Representative phospho flow overlay histograms of NF-κB p65 phosphorylation in DC subsets in PASI75 adalimumab R (blue) and NR (red). **(G)** Violin plot graphs of NF-κB p65 phosphorylation induced by various stimuli in DC subsets in PASI-75 adalimumab R (blue, n=16-20) and NR (red, n=9-10). *p* values and FDR are reported on the graph in B, D and E. **** *p* < 0.0001, *** *p* < 0.001, ** *p* < 0.01, * *p* < 0.05; Mann-Whitney U test (C, G, left and right panels) or unpaired t test (G middle panel).

Next, we sought to replicate our finding in a a second PSORT cohort of patients undergoing adalimumab therapy (PSORT adalimumab replication cohort, n=27; **Supplemental Figure 1**). Cryopreserved PBMCs were stimulated with LPS and TNF. We observed nominal correlations (all FDR>0.05) between residual disease and NF-κb phosphorylation induced by either LPS (r^2^=0.347, p=0.0209) or TNF (r^2^=0.425, p=0.0062) in cDC2, as well as TNF-induced NF-κb phosphorylation in pDC (r^2^=0.502, p=0.0021) at w0 (**Supplemental Figure 6b and Supplemental Table 4)**. Nevertheless, NF-κB phosphorylation induced by LPS and TNF in cDC2, and by TNF in pDC at w0 was significantly lower in PASI75 R than in PASI75 NR (p<0.01; **Supplemental Figure 6c**). Finally, we detected statistically significant correlations between NF-κB phosphorylation induced by either TNF (r^2^=0.285, FDR=0.029), or LPS (r^2^=0.573, FDR=10^−4^) in cDC2, as well by TNF in pDCs (r^2^=0.245, FDR=0.043) and residual disease at w0 (**Figure 2e** and **Supplemental Table 4**) in the PSORT adalimumab combined cohort (n = 43, **Figure S1**). These correlations were not driven by any clinical covariate previously associated with response to adalimumab such as age, gender, ethnicity, smoking, weight, psoriatic arthritis, being biologic naïve, the baseline PASI or the presence of the HLA-C*06:02 allele ^8,10^(**Supplemental Table 5**). In keeping with the correlation analysis, NF-κB phosphorylation at baseline was significantly lower in LPS (p<0.0001) and TNF (p<0.01)-stimulated cDC2 and TNF-stimulated pDC (p<0.0001) in PASI75 R than NR (**Figure 2f-g**), with a similar trend for most stimulations for PASI90 R and NR (**Supplemental Figure 6d)**. Interestingly, NF-κB phosphorylation induced in PASI75 R was at the same level than that observed in HV, while it was higher in NR (**Supplemental Figure 6e**). The correlation between NF-κB phosphorylation and residual disease was progressively lost in the presence of adalimumab at w 1 and w12 (**Supplemental Table 4)**, albeit TNF-(p<0.05) and LPS-(p<0.05) induced NF-κB phosphorylation in cDC2 was still significantly higher in PASI75 NR at w1 (**Supplemental Figure 6f)**.

Thus, regardless of the stimulus, NF-κB activation in cDC2 before therapy plays a role in determining clinical response to adalimumab and may have a role as predictive biomarker of response.

### Monocyte-derived dendritic cells (moDCs) of PASI75 non-responder patients display increased propensity to maturation and expression of co-stimulatory molecules at baseline

Next, we sought to gain mechanistic insights into the correlation between NF-κB activation in cDC2 cells and lack of clinical response to adalimumab. NF-κB activation drives DC maturation through the expression of co-stimulatory molecules critical for T cell priming and activation ^18^. Thus, we hypothesized that the increased NF-κB activation induced by inflammatory stimuli in cDC2 in patients failing adalimuamb therapy may reflect an increased propensity to upregulate the expression of co-stimulatory molecules.

To test our hypothesis, we generated monocyte-derived immature DCs *in vitro*, as model for cDC2, using cryopreserved baseline PBMCs of PASI75 R and NR patients (n=17) and induced DC maturation with LPS, in the presence or absence of adalimumab (**Figure 3a**). Mature moDC of PASI75 NR displayed a significantly increased expression of CD54 (p=0.0012), CD80 (p=3.11×10^−4^), CD83 (p=0.04) and CD86 (p=0.0087), and an upward trend for CD40 and HLA-DR as compared to R, with similar expression in R and HV and increased level in NR patients (**Figure 3b-c and Supplemental Figure 7a**). While addition of adalimumab significantly inhibited LPS-induced DC maturation in the overall cohort (**Supplemental Figure 7b**), the levels of CD54 (p=0.009), CD80 (p=0.003) and CD86 (p=0.02) remained significantly higher in PASI75 NR as compared to R in presence of adalimumab (**Figure 3b-c**). moDC maturation in the presence of adalimumab significantly reduced cell survival, however no difference was observed in PASI75 R and NR patients (**Supplemental Figure 7c, d**). Thus, moDC of PASI75 NR display an increased propensity to upregulate co-stimulatory molecules *in vitro* and are more refractory to the inhibitory effect exerted by adalimumab, in line with the effect exerted by adalimumab therapy on NF-kB translocation in blood DC (**Figure 1g**). The increased moDC maturation was accompanied by a significant increase and an upward trend in the production of IL12 and IL23 respectively, in the culture supernatants of mature moDC of PASI75-NR, as compared to PASI75-R, whereas there was no difference in the secretion of IL10, TNF, IL1β and IL6 (**Supplemental Figure 8)**. Taken together, lack of clinical response to adalimumab is associated with the intrinsic activation potential of cDC2 in absence of the drug.

**Figure 3.**
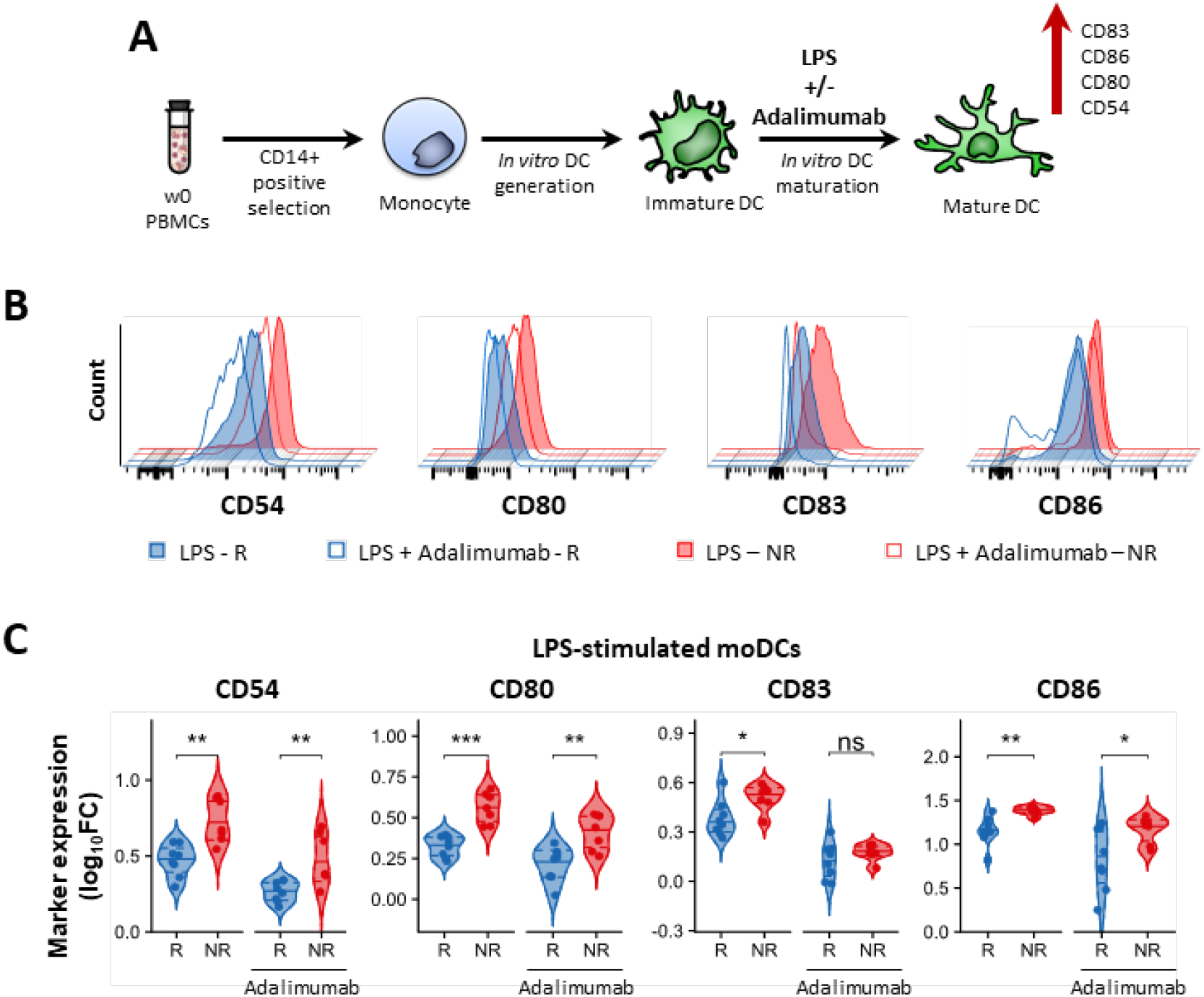
moDCs from adalimumab non-responders express higher level of co-stimulatory molecules following LPS maturation. **(A)** moDCs were generated *in vitro* from monocytes obtained from w0 PBMCs. Maturation was induced by adding LPS in the presence or absence of adalimumab. **(B)** Representative flow overlay histograms of maturation markers expression in LPS-matured (filled) and LPS + adalimumab (empty) moDCs generated in PASI75 adalimumab responders (R, blue) and non-responders (NR, red). **(C)** Violin plot graphs of maturation markers expression (measured as log10FC MFI vs Immature DCs) in PASI-75 adalimumab R (blue, n=8) and NR (red, n=7). *** *p* < 0.001, ** *p* < 0.01, * *p* < 0.05, Mann-Whitney U test (C).

### IL-17+ T cells in the blood are associated with lack of clinical reponse to adalimumab

DC maturation leads to T cell activation and cytokine production. Thus, we investigated whether the increased propensity of adalimumab NR DCs to upregulate co-stimulatory molecules *in vitro* and their resistance to the inhibitory effect of adalimumab (**Figure 3, Figure 1g**) has an effect on T cell responses and can be detected in circulating DCs. To this end, we studied the phenotype of T cells and DCs in the blood of psoriasis patients before and during adalimumab therapy.

First, we studied circulating T cells producing the psoriasis hallmark cytokine IL-17A alonside IL-10 producing T regulatory (Treg) in psoriasis patients and HV (**Supplemental Figure 9**). As previously reported ^19^, frequency of IL17A^+^CD4^+^T (Th17), IL17A^+^CD8^+^ T (Tc17) and IL-17A^+^Treg cells was significantly increased at baseline in PBMCs of psoriasis patients as compared to healthy volunteers (**Supplemental Figure 10)**. Tc17 and IL-17A^+^Treg cells significantly decreased during adalimumab therapy, while frequency of Th17 and IL10^+^Treg cells did not change (**Figure 4a**). Moreover, we detected a statistically significant correlation between residual disease and the frequency of Th17 and IL-17A^+^Treg at w0 and of Tc17 at w12 (**Figure 4b and Supplemental Table 6**). In keeping with this finding, PASI75 NR had significantly higher frequency of Th17 and IL-17A+Treg at w0 and Tc17 at week 12 (**Figure 4c**). Moreover, frequency of Th17 and IL-17A+Tregs cells was similar in HV and PASI75 R, but significantly higher in NR (**Supplemental Figure 10b**). On the other hand, frequency of Tc17 cells at w0 was significantly higher in both R and NR, but decreased to HV level in PASI75 R at w12. Thus, clinical response to adalimumab is associated with a decrease in circulating Tc17 at week 12, in keeping with the emerging critical role for CD8+ T cells in psoriasis^20,21^. Moreover, frequency of Th17 cells prior to therapy may have a role as predictive biomarker of response, in line with the heightened NF-kB activation and maturation status induced in DCs.

**Figure 4.**
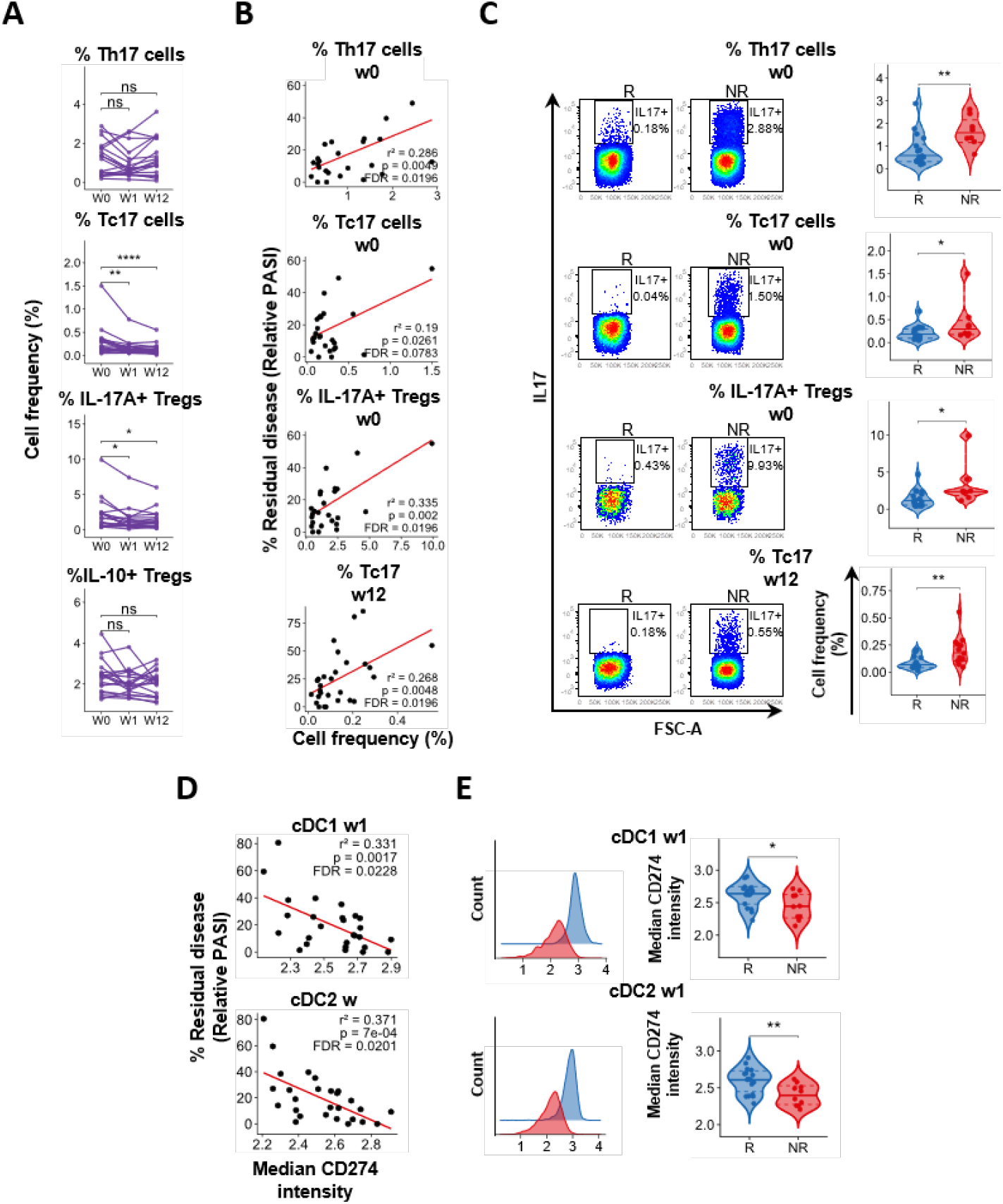
Lack of response to adalimumab correlates with increased levels of blood IL17A+ T cells at w0 and increased expression of CD274 in DC at w1. Immunophenotyping of PBMCs of psoriasis patients at w0 and at w1 and w12 after starting adalimumab therapy. **(A)** Frequency of IL10+ and IL17A+ CD4, CD8 T cells and Tregs at different time points, each line represents one patient (n=19). **(B)** Correlation analyses between the frequency of IL17A+ or IL10+ cells within CD4, CD8 or Tregs and residual disease at w12 (n=26-28). **(C)** Representative flow cytometry colour-plots (left) and violin plots graphs of the frequency of cytokine-producing cells in adalimumab PASI75 responders (R, blue, n=15-18) and non-responders (NR, red, n=8-13). **(D)** Correlation analyses between the expression of CD274 at w1 and residual disease at w12 in cDC1 and cDC2 (n=27). **(E)** Representative histograms overlay graphs (left) and violin plots graphs (right) of CD274 expression in DC of PASI75 adalimumab R (blue, n=17) and NR (red, n=10). **** *p* < 0.0001, ** *p* < 0.001, ** *p* < 0.01, * *p* < 0.05, Kruskal–Wallis with Dunn’s multiple comparisons post-test (A), Mann-Whitney U test (C) or unpaired t test (E).

Next, we deeply phenotyped blood DC subsets (Combined PSORT adalimumab cohort, n=43) using a 15-colours flow-cytometry panel comprehensive of activation (HLA-DR, CCR7, CD40), maturation (CD80, CD83, CD86), adhesion (CD54, CD209) and inhibitory (CD274) markers, and applied Flow self-organizing map (SOM) unsupervised clustering analysis ^22,23^ to identify cell populations and DC subsets **(Supplemental Figure 11a)**. Frequency of cDC1 was significantly decreased at week 12 of adalimumab therapy, while cDC2 and pDc were unaffected (**Supplemental Figure 11b**) but neither of them correlated at any time point with clinical response at week 12 (**Supplemental Figure 11c**). Overall, marker expression did not change over time (**Supplemental Figure 12 and Supplemental Table 7**), or correlate, at any time point, with clinical response at week 12 (**Supplemental Table 8**) or differ between PASI75 R and NR (**Supplemental Table 9**). Nevertheless, we detected statistically significant correlations between relative PASI and CD274 expression at week 1 in cDC1 (r^2^=0.331, FDR<0.05) and cDC2 (r^2^=0.371, FDR<0.05) (**Figure 4d** and **Supplemental Table 8**) and CD274 expression in cDC1 (p<0.05) and cDC2 (p<0.01) at week 1 was nominally different (FDR>0.05) in PASI75 R and NR (**Figure 4e and Supplemental Table 9**).

To explore a possible genetic influence on *CD274* expression, we mined the Genotype-Tissue Expression database (GTEX, www.gtexportal.org) and identified rs59906468 as a strong cis eQTL (beta = 0.26, p=3.6×10^−25^) in whole blood, with the A allele reducing the expression of *CD274* (**Supplemental Figure 13a**). We observed a similar trend in our adalimumab cohort for CD274 protein expression in DC subsets at various time points (**Supplemental Figure 13b**). Moreover, in an extended PSORT genetic dataset ^10^, several putative associations with adalimumab response were observed in the CD274 region at both 3m and 6m for both PASI75 and PASI90 response outcomes (p-values from 0.10 to 8.8×10^−6^; **Supplemental Figure 13c**). Bayesian colocalization analysis suggested that rs59906468 may be a shared causal variant for both *CD274* expression and adalimumab response, particularly PASI90 response at 6m (posterior probability for shared effect = 0.66; **Supplemental Figure 13d**).

Thus, deep phenotyping of blood DCs suggests that clinical response to adalimumab is associated with early upregulation of the inhibitory molecule CD274 in DCs, which may be genetically driven. However, the surface phenotype of blood DCs prior to therapy does not play a major role in determining clinical response nor has biomarker potential.

### Psorasis skin of PASI75 non-responders has the hallmarks of increased DC maturation and T17 activation before adalimumab therapy

DC maturation is triggered at sites of inflammation and mature DCs are abundant in psoriasis lesions. Thus, the heightened NF-kB activation and maturation propensity detected in blood DCs following *ex vivo* and *in vitro* stimulation may have an effect on the surface phenoptye of skin rather than circulating DCs. To this end, we studied the effect of adalimumab on DC, as well as, T cell phenotype in lesional skin sections of PASI75 R and NR at w0 and w12 (n=20). Adalimumab therapy significantly decreased the number of CD11c+ DC (p<0.01) and their expression of CD83 (p<0.01), as well as the number of IL23+ (p<0.01) DC at week 12 (**Figure 5a and Supplemental Figure 14**). Moreover, PASI75-NR harboured increased number (p<0.05) of CD11c+ DC which had enhanced expression of CD83 (p<0.05) and produced IL23 in greater number (p<0.05) as compared to PASI75-R at w0 (**Figure 5b,c**), confirming a more mature skin DC phenotype in NR, in line with the DC maturation data obtained *in vitro* (**Figure 3)**. While PDL-1 expression in CD11c DC increased with therapy in the overall cohort (**Supplemental Figure 14b)**, no difference was observed between R and NR at any time point (**Supplemental Figure 14c-d**). As expected, the number of IL17+ CD4+ (Th17, p<0.01) and CD8+ T (Tc17, p<0.05) cells present in psoriatic skin at w12 (**Figure 5d**) was decreased by adalimumab therapy. The number of Th17 (p<0.05) and Tc17 (p<0.05) cells was significantly higher in the skin of NR at w0 (**Figure 5f,g**). Of interest, while number of Th17 at w12 was similar in R and NR, Tc17 cells remained significantly elevated (p<0.01) in the skin of NR as compared to R (**Figure 5e,f**), in line with the association between clinical response to adalimumab and the decrease in circulating Tc17 cells observed in blood at w12 (**Figure 4**).

**Figure 5.**
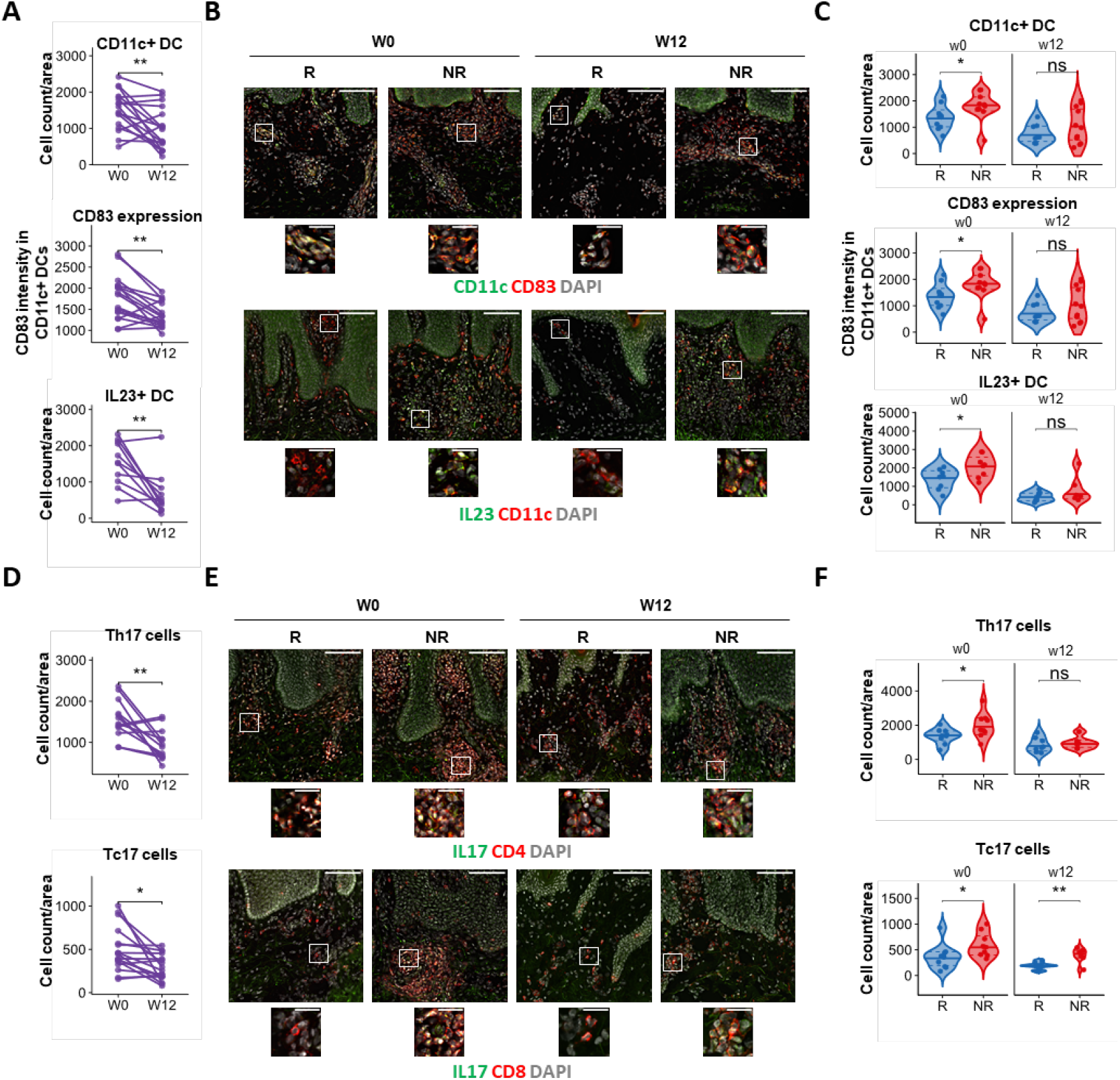
Lack of response to adalimumab is associated with increased DC and T17 activation before therapy. Analysis of psoriasis lesional skin of patients at baseline (w0) and after 12 weeks (w12) of adalimumab treatment **(A)** Number of CD11c+ DCs, expression of CD83 in CD11c+ dermal dendritic cells (DC) and number of IL-23+CD11c+ DC per area of analysis (750×750μm) at different time points. Each line represents one patient. **(B)** Representative immunofluorescence images of psoriasis skin showing total number of CD11c+ DCs (top), CD83 expression in CD11c+ DC (middle) and IL23+CD11c+DC (bottom) in PASI75 adalimumab responders (R) and non-responders (NR). **(C)** Violin plot graphs of CD11c+ DC counts (top), CD83 expression in dermal DC (middle) and number of IL23+DC in adalimumab R (blue, n=8-10) and NR (red, n=6-10) at w0 and w12). **(D)** Number of IL-17A+CD4+ Th17 and IL-17A+CD8+ Tc17 cells per area of analysis at different time points. Each line represents one patient. **(E)** Representative immunofluorescence images of psoriasis skin showing Th17 (top) and Tc17 (bottom) in adalimumab R and NR. **(F)** Violin plot graphs of number of Th17 and Tc17 cells in lesional skin sections of in adalimumab R (blue, n=8-10) and NR (red, n=6-9) at w0 and w12. ** *p* < 0.01, * *p* < 0.05, Wilcoxon test (A bottom panel), or paired t test (A minus bottom panel, D), Mann Whitney-U test (C top panel, F) or unpaired t test (C minus top panel). Scale bars in B, E: 100 μm (overview) and 25 μm (insets).

### NF-κB phosphorylation in cDC2 before therapy is a predictive biomarker of response to adalimumab

Finally, we evaluated the predictive value for patient stratification of the 8 blood immune traits at w0 or w1 which significantly correlated with clinical response at w12 (i.e. NF-κB phosphorylation at w0 induced by LPS in cDC2 and by TNF in pDc and cDC2; frequency of Th17, Tc17 and IL-17+ Tregs at w0, CD274 expression at w1 in cDC1 and cDC2). We built receiver operating characteristic (ROC) curves for the binary outcome of PASI75 R or NR at w12 for each potential biological read-out (NF-κB phosphorylation, T cell frequency, CD274 expression) obtained in the combined cohort (**Supplemental Figure 15a**) and ranked them according to their area under the curve (AUC). The top markers were: LPS-induced NF-κB phosphorylation in cDC2 at w0, frequency of Th17 at w0 and expression of CD274 in cDC2 at w1, with an AUC of 0.922, 0.840 and 0.806, respectively **(Supplemental Figure 15a and Figure 6a**). The best performing biomarker overall, NF-κB phosphorylation in cDC2 (cut-off threshold of 0.169 log_10_FC) had 80 % sensitivity, 88.9 %specificity, 85.7% accuracy, and predictive odds ratio of 32 (**Figure 6b**). AUC did not improve when we combined LPS-induced NF-κB phosphorylation in cDC2 with the other top signals identified (**Figure 6c**), therefore we selected this biomarker for further investigation and validation. The psoriasis-susceptibility HLA-C*06:02 allele has been associated with clinical response to biologics, with carriers less likely to respond to adalimumab ^10^. However, inclusion of HLA-C*06:02 genotype in our models did not increase the predicitve value of LPS-induced NF-κB phosphorylation in cDC2 (**Figure 6d**). Follow-up data for 26 of the 28 patients assessed at w12, showed that LPS-induced NF-κB phosphorylation in cDC2 at w0 also predicted PASI75 response at 6 months (cut-off threshold 0.151 log_10_FC, 85.7 % sensitivity, 73.7 % specificity and AUC of 0.80 (p<0.05) (**Supplemental Figure 15b**). Finally, we tested the ability of the model to predict clinical response to adalimumab at w12 in an additional independent cohort of 15 psoriasis patients (Clinical validation cohort, **Supplemental Figure 1**). Applying the cut-off threshold of 0.169 log_10_FC for LPS-induced NF-κB phosphorylation in cDC2, the model predicted treatment outcome to adalimumab at w12 with 93.3% accuracy, 100 % sensitivity 90.1 % specificity (**Figure 6e**).

**Figure 6.**
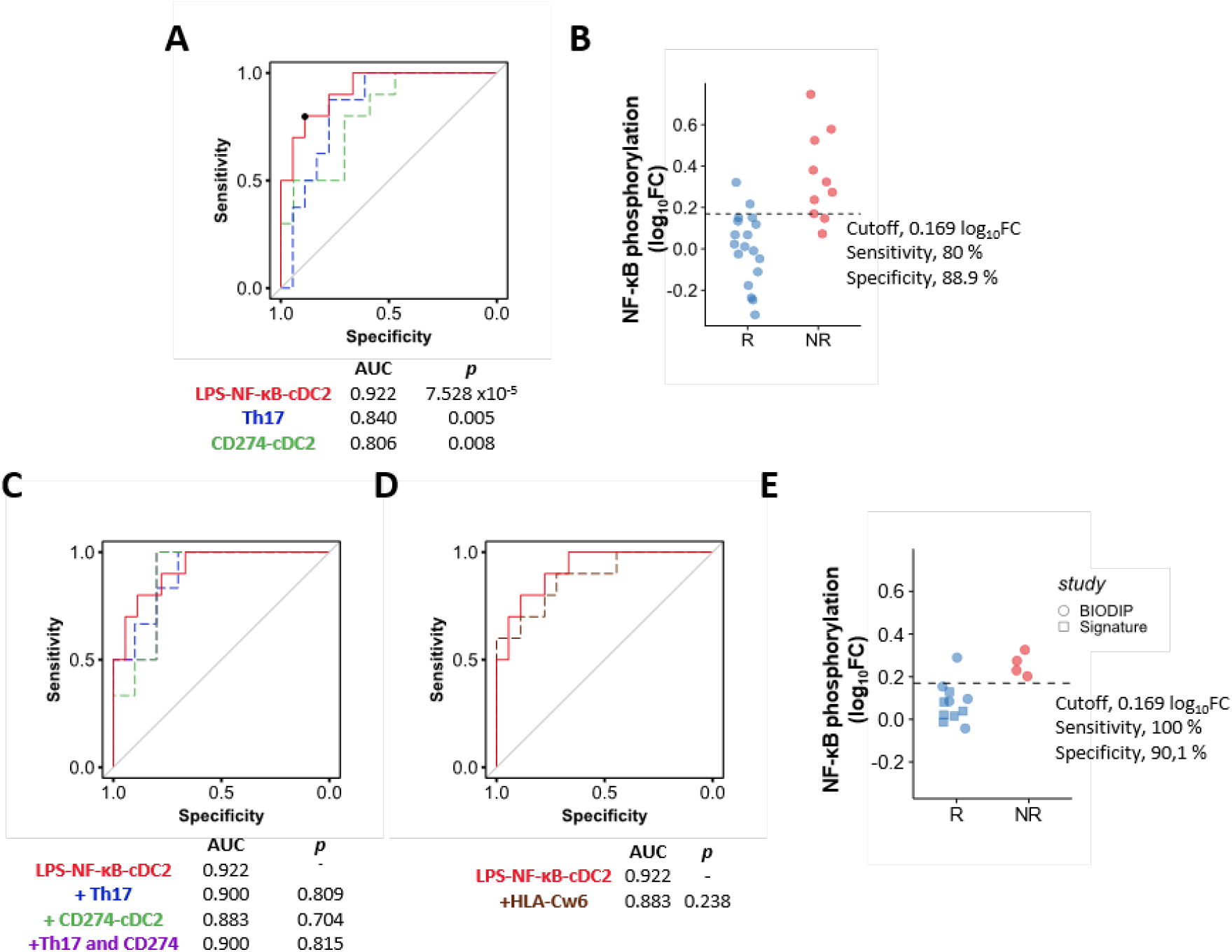
NF-κB phosphorylation in cDC2 at baseline is a predictive biomarker of response to adalimumab at week 12. **(A)** Receiver operator characteristic (ROC) of linear models for LPS-induced NF-κB phosphorylation in cDC2 at baseline (w0, red line), frequency of blood Th17 cells at baseline (w0, blue line) and PDL1 expression in cDC2 at w1 (green line) predicting PASI75 outcome in adalimumab patients at week 12 (combined cohort). Black dot shows the cut-off for maximal accuracy for the best ROC curve. p values *vs* an AUC of 0.5 are shown on the graph. **(B)** Dot plot graph of the PSORT adalimumab combined cohort (n = 28) classified in PASI75 responder (R, blue, n=18) and non-responders (NR, red, n=10) according to the cut-off for LPS-induced NF-κB phosphorylation in cDC2 at w0. **(C)** ROC of LPS-induced NF-κB phosphorylation in cDC2 at w0 (solid red line) and combination biomarkers (LPS-NF-kB-cDC2 + Th17, dotted blue line; LPS-NF-kB-cDC2 + CD274-cDC2 dotted green line, and LPS-NF-kB-cDC2 + Th17 + CD274-DC2, dotted purple line). The purple line partially overlaps with the blue line. p values *vs* AUC of LPS-NF-kB-cDC2 are shown. **(D)** ROC analysis for LPS-NF-kB-cDC2 predicting PASI75 outcome in adalimumab patients (solid red line) and when combined with HLA-C*06:02 genotype (dotted brown). p value vs AUC of LPS-NF-kB-cDC2 is shown. **(E)** ROC clinical validation using optimal cut-off for LPS-induced NF-κB phosphorylation in cDC2 at w0 to classify an independent cohort of adalimumab patient in PASI75 R (blue, n=11) and NR (red, n=4) (clinical validation cohort). Two-tailed Mann–Whitney U test, A, C, D.

Taken together, our results clearly identify TNF and LPS-induced NF-κB phosphorylation in cDC2 dendritic cells prior to therapy as a predictive biomarker of response to adalimumab which may aid in patient stratification.

## Discussion

Here we report the discovery, replication, and mechanistic investigation of an actionable, predictive biomarker of response to the anti-TNF adalimumab in psoriasis. NF-κB activation of blood cDC2, measured as LPS-induced NF-κBp65 phosphorylation before therapy, correlated with clinical outcome, discriminating between PASI R and NR with high sensitivity and specificity for up to 6 months after commencing therapy. The increased induced NF-kB activation detected in NR before commencing adalimumab was mechanistically linked to increased DC maturation and resistance to the inhibitory effect of adalimumab *in vitro*, as well as well increased T17 cell activation in blood, ultimately resulting in a more mature and proinflammatory DC phenotype and increased number of T17 cells in the skin.

NF-κB signaling plays a critical role in inflammation and multiple genes of this pathway are associated with psoriasis susceptibility ^24^. Activation of NF-κB by inflammatory stimuli, through engagement of TLRs and cytokine receptors, culminates in the translocation of NF-κB complexes into the nucleus and activation of gene expression. Phosphorylation of p65 NF-κB subunit, such as at Ser529, further regulate NF-κB activation and function by increasing its transcriptional activity ^17^. We show that *ex vivo* TNF-induced NF-κB translocation was largely inhibited in most immune cell types by the free drug present in the blood of patients sampled during therapy. Nevertheless, the near complete inhibition of induced NF-κB signaling in T cells mediated by adalimumab does not underpin its mechanism of action in psoriasis as it deos not correlate with clinical response. On the other hand, TNF-induced NF-kB activation in DC was only partially inhibited by adalimumab, with DC from NR more refractory to its inhibitory effect. Moreover, NF-κB activation induced in DC by various pro-inflammatory stimuli before commencing therapy, and measured as either nuclear translocation or p65 phosphorylation, correlated with lack of clinical response at w12.

DC are highly implicated in psoriasis pathogenesis ^2,3^. Mature myeloid cDC with high T-cell stimulatory capacity are enriched in skin lesions where they produce IL-23 and other proinflammatory cytokines ^25^ and are poised to present putative auto-antigens, such as LL37, to T cells. Activation of NF-κB signaling in DC, triggered by either pathogen-associated molecular patterns or proinflammatory cytokines, induces most of the phenotypic and functional characteristics typical of mature DCs, inducing the expression of MHC II and co-stimulatory molecules ^26,27^. TNF blockade by anti-TNF etanercept and infliximab impairs maturation of DC generated *in vitro* from healthy volunteers, resulting in reduced levels of HLA-DR and co-stimulatory molecules ^16,28^. Moreover, the expression of co-stimulatory molecules in DCs from psoriasis skin diminished during etanercept therapy ^16^ and *in vitro*-derived DC from rheumatoid arthritis patients receiving anti-TNF displayed impaired upregulation of co-stimulatory molecules and poor T-cell stimulatory activity ^28^. Our data build on these previous findings, validating cDC2 as the key mechanistic cellular target of adalimumab influencing clinical outcome. In keeping with the increased NF-κB activation, the phenotype of *in vitro-*generated DC in PASI75-NR before commencing adalimumab was strikingly different from DC generated from R and healthy volunteers. NR DC displayed a more mature phenotype, which was only partially inhibited by *in vitro* adalimumab, suggesting that an intrinsic increased propensity to respond to inflammatory and maturation stimuli limits the beneficial effect of TNF-blockade. Crucially, before commencing the therapy, the skin of PASI75 NR patients harbored DC with a more mature and activated phenotype, thus validating the *in vitro* findings. Moreover, the lack of phenotypic differences in circulating blood DC prior to therapy suggests that a pro-inflammatory environment is required to trigger such differences, otherwise undiscernible in resting *ex vivo* cells.

The significant correlation between clinical response and NF-κBp65 phosphorylation in cDC2, induced by TNF or LPS indicates a mechanistic link between DC intrinsic maturation potential and adalimumab clinical efficacy. Not surprisingly, the correlation with the best predictive value was detected in LPS-stimulated cells, as LPS is more efficient than TNF to mature DCs *in vitro* ^29^. Nevertheless, LPS induces autocrine TNF in DC ^30^, thus providing a further biological rationale between the identified biomarker and the biological pathway targeted by adalimumab.

DC immunogenicity is determined both by their maturation state and their lifespan. moDCs and blood cDC2 express both TNF receptor (TNFR)-1 and TNFR2, withTNFR1 the major TNFR controlling their maturation through the integrated activation of “canonical” NF-κB p65 and “non canonical” NF-κB RelB pathway ^29,31^. Moreover, p65 NF-κB mediates the pro-survival effect mediated by TNFR1 in DC ^29^. It is possible that DC of NR patients may also have increased survival and extended lifespan, as suggested by the induction of apoptosis detected during DC maturation in the presence of *in vitro* TNF blockade ^28^. However, conflicting data have been observed in psoriasis skin ^14,32^. In our *in vitro* system, adalimumab significantly decreased cell viability measured at day 8 of culture, with a minimal upward trend detected in NR patients. Similarly, although the overall number of CD11c+ DC was decreased at w12, it did not discriminate between PASI75 R and NR, suggesting that, although affected by TNF-blockade, DC survival is unlikely to determine clinical response to adalimumab.

An intriguing possibility is that the expression of the immune checkpoint CD274/PDL-1 may be involved in underpinning DC as cellular determinants of response to adalimumab, as suggested by the correlation between CD274 expression and clinical response. Moreover, this effect may be genetically driven with at least one eQTL possibly explaining both CD274 expression and response to adalimumab. CD274, expressed by DC, is the main inhibitory ligand of CD279, also known as PD-1, expressed on T cells. Engagement of CD279 by CD274 alters T cell activity, by inhibiting T cell proliferation, survival and cytokine production ^33^. CD279/PD1 is expressed on IL-17A+ T cells in psoriasis lesions and its blockade ameliorates inflammation in the aldara-induced psoriasiform skin inflammation model ^34^. Another link between CD274/PDL-1 and psoriasis stems from the clinical observation than more than one third of cancer patients treated with Immune checkpoint inhibitors targeting cytotoxic T lymphocyte-associated antigen-4 (CTLA-4), CD279/PD-1 or CD274/PD-L1 develop cutaneous immune-related adverse events (irAEs), including characteristic psoriasis lesions ^35^.

Early studies have investigated the effect of TNF blockade on circulating 17A-producing T cells, with conflicting results. Two studies reported only a modest and insignificant decrease in frequency of circulating Th17 cells in patients receiving any of the anti-TNF agents ^19,36^, while another two found a significant decrease in circulating Th17 cells in patients undergoing adalimumab or etanercept treatment ^37,38^. We found that frequency of circulating Th17 cells did not change during adalimumab treatment, while circulating Tc17 cells dramatically decreased. More importantly, frequency of IL17A+ T cells, either Th, Tc or Treg, was higher in PASI75 NR before commencing adalimumab, while Tc17 remained increased in PASI75NR at week12. An association between high baseline frequency of Th17 and lack of response to anti-TNFs has also been reported in RA ^39^. In keeping with our blood data, the number of both Th17 and Tc17 cells in lesional psoriasis skin was significantly higher in PASI75 NR at w0, but only Tc17 cells remained higher in NR at w12. Thus, while frequency of circulating T17 before the start of the therapy may have a role in determining clinical outcome, an effective clinical response is only achieved in the presence of significant decrease of Tc17 cells in both blood and skin. This is in line with the retainment of IL-17+CD8+ Tissue-resident memory (T_rm_) cells in clinically resolved psoriasis lesion, where they are poised to be reactivated ^40^, and the beneficial effect of CD8 blockade in preventing psoriasis development in the AGR 129 xenotransplant model ^21^. Nevertheless, our data show that despite the total inhibition of NF-kB signaling in T cells exerted by adalimumab, clinical response is determined by its effect on DC, which in turn affects T cells in blood and skin.

Taken together, our study uncovers, for the first time, key baseline determinants of response to adalimumab in psoriasis at molecular and cellular levels, from the more proximal signaling events, such as NF-κB activation in cDC2, to the ultimate effect on pathogenic T17 cells in blood and skin. Adalimumab is less effective in individuals in which increased NF-κB signaling induces a heightened maturation phenotype in DC, which in turns results in increased frequency of T17 cells in blood and skin before the start of the therapy (**Figure 7)**. These findings suggest the existence of a specific disease endotype with direct implications on clinical response to adalimumab. Importantly, we identified, replicated and mechanistically validated LPS-induced NF-κB p65 phosphorylation in cDC2 at baseline as a predictive biomarker of clinical response to adalimumab. Blood biomarkers are minimally invasive and transcription factor phosphorylation is already a validated diagnostic tool in cancer ^41^, thus our essay is easily scalable for further clinical validation in future prospective clinical trials. Importantly, such trials should also address the predictive value of the test not only for other anti-TNF agents but particularly for their more recent cheaper biosimilars ^42^, providing an alternative to the more expensive biologics targeting the IL-23/IL-17 pathway. Finally, our findings have the potential to inform patient stratification in other immune-mediated inflammatory diseases such as inflammatory bowel disease and rheumatoid arthritis where TNF-blockade is also a therapeutic cornerstone ^39,43^.

**Figure 7.**
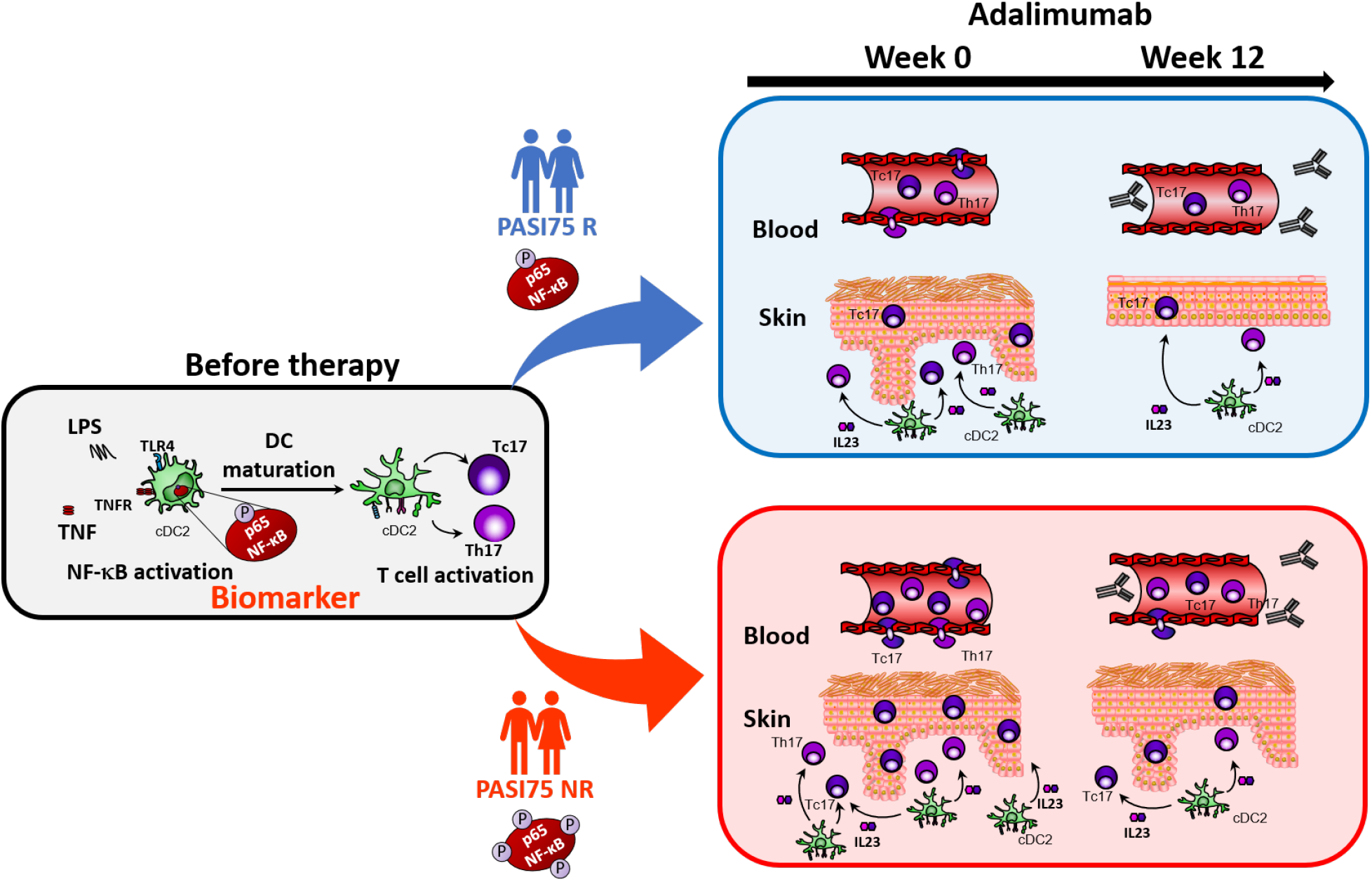
Mechanism of action of adalimumab and predictive biomarker of clinical response. Phosphorylation of p65NF-κB in conventional Type 2 dendritic cells (cDC2) before therapy is an early predictive biomarker of response to adalimumab at week 12. NF-kB activation by TNF or TLR ligands, e.g LPS, induces DC maturation which in turn leads to the activation of IL-17A producing T helper (Th)17 and CD8+ T cytotoxic (Tc)17 cells. This is enhanced in PASI 75 non responder patients (PASI75 NR), as shown in the cartoon by the increased number of phosphorylation sites on NF-kB underneath the patient silhouettes, and results in increased number of activated DC producing IL-23 in the skin and of Th17 and Tc17 in blood and skin before therapy, as compared to PASI75 responders (PASI75 R). TNF-blockade by adalimumab reduces DC and T cell responses but they remain elevated in the blood and skin of PASI75 NR underpinning the lack of clinical response.

## Materials and Methods

### Study design and patient cohorts

The study included 67 adult patients with chronic plaque type psoriasis with moderate-severe disease (PASI>10) recruited into the Psoriasis Stratification to Optimise Relevant Therapy (PSORT) study ^7^ at 6 centers in the UK between May 2015 and July 2018 and due to start biologic therapy (ustekinumab or adalimumab) as part of routine clinical practice (**Supplemental Table 10**). Exclusion criteria were the use of systemic or biological therapy for psoriasis for 2 weeks prior to study entry, use of PUVA therapy for 3 months or UV-B for 1 month prior to study entry or use of topical treatments to site of biopsies (except for emollients) for 2 weeks prior to study entry, as well as serious/uncontrolled systemic disease or medical condition. The discovery cohort includes 16 PSORT patients receiving adalimumab and 24 patients receiving ustekinumab, the replication cohort includes a further 27 PSORT patients receiving adalimumab; all 43 PSORT patients receiving adalimumab constitutes the combined adalimumab cohort (**Supplemental Figure 1, Supplemental Table 10**). Patients were largely of Caucasian descent, both genders were represented (29 males/14 females), age range was 22–72 years, mean 45 years. The clinical validation cohort **(Supplemental Figure 1)** comprises of additional 15 psoriasis patients not receiving biological treatment at time of sampling. This included 9 patients prospectively recruited into the BIODIP study ^44^ to receive adalimumab, sampled at baseline (w0) before commencing therapy, and 6 patients recruited into the Signature study ^45^, who had previously received adalimumab and had reached PASI75 at w12, but had then progressively lost clinical response 10-41 months after starting therapy and were retrospectively sampled 1-11 months after stopping adalimumab and before commencing secukinumab. Thus, for the purpose of our analysis they were considered PASI75 R at w12.

This study was performed in accordance with the declaration of Helsinki and was approved by the London Bridge research ethics committee (REC numbers: 14/LO/1685; 11/LO/1692; 06/Q0704/18; 13/EE/0241). All participants provided written informed consent.

Full clinical and demographics patient information are in **Supplemental Table 10**.

Samples were randomly assigned to experiment batches, unless otherwise stated, and blinded for the person performing the experiments and the analyst carrying manual analysis steps such as cell gating. Some samples were excluded from the analysis following quality control (QC).

### Blood samples and PBMCs isolation

Blood was collected at participating sites in BD Vacutainer™ Hemogard Closure Plastic K2-Edta Tubes (BD Biosciences, CA). For the discovery cohort, an aliquot of fresh blood was used for imaging flow cytometry (see below). Peripheral blood mononuclear cells (PBMCs) were isolated within 4 hours of collection using Ficoll-Paque density gradient centrifugation in Leucosep tubes (Greiner Bio-One, Austria). Cells were viably cryopreserved as previously described ^44^ and stored in liquid nitrogen until used or shipped to St John’s Institute of Dermatology. Prior to each experiment, cell count and viability were measured with Via1-Cassette™ on a NucleoCounter NC-200 (Chemometec, DK).

### Skin samples

6mm punch lesional psoriasis skin biopsies were obtained from patients and bisected. Half was embedded in optimal cutting temperature compound (OCT; VWR, PA) and stored in liquid nitrogen until frozen-sectioned on a microtome-cryostat.

### Imaging flow cytometry

200 μl of fresh whole blood were incubated with Fc block (BioLegend, CA) for 10 min at room temperature (RT). Surface staining antibodies (Imaging flow cytometry panel, **Supplemental Table 11** and **Supplemental Figure 2**) were added and cells were stimulated with either TNF 20 ng/ml or LPS 1 μg/ml (Sigma-Aldrich, UK) or left unstimulated for 30 min at 37 °C. In some experiments, adalimumab 10 μg/ml (AbbVie, IL), which approximates adalimumab plasma concentration when given 40 mg biweekly^9^, was added 30 minutes before stimulation. Erythrocytes were lysed and samples were simultaneously fixed with BD Phosflow™ Lyse/Fix Buffer (BD Biosciences, CA) for 10 min at 37 °C. Cells were spun down and intracellular staining primary rabbit NF-κB p65 (Cell Signalling, MA) (Imaging flow cytometry panel, **Supplemental Table 11**) was added in permeabilization buffer (0.1% Triton X in PBS) and incubated for 20 min at RT. Cells were washed in 2% BSA, 1mM EDTA PBS and intracellular staining secondary donkey anti-rabbit IgG (BioLegend, CA)(**Supplemental Table 11**) was added in permeabilization buffer and incubated for 20 min at room temperature. Samples were washed and DAPI (Invitrogen, CA) 25 ng/ml was added prior to acquisition on an ImageStream™ MarkII imaging cytometer (Amnis, WA) at X60 magnification. Gating of cell populations of interest and image analysis was performed using the IDEAS software (Amnis, WA). Nuclear translocation was assessed by the median internalization score feature for NF-κB signal in the nuclear mask based on the DAPI staining, which represents constitutive nuclear localization. The relative shift in the distribution between unstimulated *versus* stimulated sample was calculated using Fisher’s Discriminant ratio (R_d_ score) according to the formula:

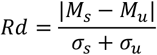

Cell populations for which less than 10 cells were acquired and/or stimulation conditions in which the induced R_d_ score was lower than 0.3 were excluded from the analysis.

### Phospho flow cytometry

Cryopreserved PBMCs were thawed in 22 batches of 1-2 patients per experiments, with each experiment comprising all time points (visits). Cells were rested overnight in RPMI-1640 medium supplemented with 10% heat-inactivated new-born calf serum (Life Technologies, CA) and penicillin/streptavidin 1%. Next day, 2×10^6^ cells were stimulated with either TNF 100 ng/ml, IL1β 10 ng/ml (PeproTech, UK) or LPS 1 μg/ml (Sigma-Aldrich, UK) for 15 minutes at 37 °C. Samples were placed on ice and surface staining antibodies in brilliant stain buffer (BD Biosciences, CA) (Phospho Flow Cytometry panel, **Supplemental Table 11** and **Supplemental Figure 4**) were added for 30 minutes. Subsequent steps were carried out at RT. Cells were fixed with 1.5% paraformaldehyde in deionized water (Electron Microscopy Science, PA) for 10 minutes, permeabilized with Perm Buffer IV (BD Biosciences, NJ) for 20 minutes and stained with intracellular PE NF-κB p-p65 (Ser529) antibody (BD Biosciences, CA) for 1 hour (**Supplemental Table 11**). Samples were acquired on a BD LSRFortessa™. NF-kB p65 Ser529 phosphorylation was calculated as log_10_ Mean fluorescence intensity (MFI) of stimulated sample /MFI of unstimulated sample, abbreviated as log_10_ fold change (FC) MFI. QC of the samples based on cell viability was carried out by building a receiver operating characteristic (ROC) curve to identify viability cut-off which best discriminate good quality samples, pre-set as those with CD14+ cell frequency >5% and LPS-induced NF-κB phosphorylation >0.2log_10_FC (**Supplemental Figure 16**). As a result, samples with cell viability <93% (sensitivity 90% and specificity 86%) were not included in the analysis. Inter-experiment variation in terms of cell frequency and level of NF-κB phosphorylation was assessed by running one internal control (IC) sample in each experiment. Inter-experiment variation for IC resulted to be minimal, while variability among patient samples was much larger, as expected for true biological variation (**Supplemental Figure 17**).

### Monocyte-derived dendritic cell (moDC) generation, maturation and phenotyping

Cryopreserved psoriasis patient PBMCs collected at w0 were thawed in 5 batches of 4 patients per experiment. Monocytes were isolated from PBMCs using MACS Separation Columns and CD14 MicroBeads (Miltenyi, Germany) as described by the manufacturer, and their purity was checked by flow cytometry (average purity > 99%, **Supplemental Figure 18**). Cells were cultured in RPMI-1640 medium supplemented with 10% heat-inactivated new-born calf serum (Life Technologies, CA), penicillin/streptavidin 1% for 6 days in the presence of IL-4 (500 U/ml) and GM-CSF (1000 U/ml) (PeproTech, UK) to obtain monocyte derived dendritic cells (moDC), as described previously ^46^. DC maturation was induced by adding LPS 250 ng/ml (Sigma-Aldrich, UK) in the presence or absence of adalimumab 10 ug/ml (AbbVie, IL) on day 6 of culture. On day 8, cells were stained with antibodies of the moDC panel (**Supplemental Table 11**) in brilliant stain buffer (BD Biosciences, CA) (Supp table 13) for 30 minutes on ice, fixed with 1.5% paraformaldehyde (Electron Microscopy Science, PA) for 10 minutes and acquired on a BD FACSCanto™. Samples with viability < 85% were excluded from the analysis. Marker expression was reported as log_10_ MFI of LPS-matured moDC /MFI of immature moDC, abbreviated as log_10_ FC MFI. Experiments in healthy volunteer were run according to the same protocol by a different operator at a later time. To avoid operator-depended batch effects, a validation experiment was run with 2 samples each from psoriasis adalimumab PASI75 R, PASI75 NR and HV and a normalization factor was calculated by dividing the median log_10_FC MFI of the psoriasis patient experiments and the healthy volunteer experiments by the median of these groups obtained in the validation experiment. Thus, normalized marker expression is reported in **Supplemental Figure 7** comparing patients and HV. Cytokine levels in day 8 moDC culture supernatant were assessed using Milliplex Map Human TH17 Magnetic Bead Panel (Merck Millipore, MA), following manufacturer’s instructions.

### T cell phenotyping

Cryopreserved PBMCs were thawed in 13 batches of 3-4 patients per experiments, with each experiment comprising w0 and w12 visits of interest. Cells were rested overnight in RPMI-1640 medium supplemented with 10% heat-inactivated new-born calf serum (Life Technologies, CA) and penicillin/streptavidin 1%. Next day 3×10^6^ cells were stimulated with PMA (50 ng/ml, Sigma-Aldrich, UK) and ionomycin (1 mg/ml, Sigma-Aldrich, UK), in the presence of monensin (2 μM, Invitrogen, CA) and brefeldin A (3 μg/ml, Invitrogen, CA) for 3 hours at 37 °C. Then, cells were centrifuged, resuspended in PBS and incubated with live/dead FVS780 reagent (BD Biosciences, CA) on ice for 30 min. Cells were spun down and stained with surface antibodies in brilliant stain buffer (BD Biosciences, CA) of the T cell phenotyping panel **(Supplemental Table 11**) for 30 minutes on ice, fixed and permeabilized with the Human FoxP3 Buffer Set (BD Biosciences, CA) following manufacturer’s instructions. Intracellular antibodies were added for 1 hour, cells were washed and acquired on a BD FACSCanto™. Samples with viability < 85% were excluded from the analysis. Inter-experiment variation in terms of cell frequency (**Supplemental Figure 19a**) and cytokine production (**Supplemental Figure 19b**) was assessed by running one IC sample in each experiment. Inter-experiment variation for IC resulted to be minimal, while variability among patient samples was significantly larger, as expected for true biological variation (**Supplemental Figure 19**)

### Flow cytometry data acquisition, pre-processing and analysis

Flow and phospho flow cytometry data were acquired using BD Standardized Application Setup, which allows to combine data acquired in different experiments thus minimizing batch-effects, and data were compensated using FACSDiva software. Signal anomalies derived from abrupt changes in the flow rate, instability of signal acquisition and/or margin events in the lower or upper limit of the dynamic range were filtered out of individual sample fcs files with the R package flowAI ^47^. Cells were manually gated using Flowjo™ software 10.6.1. A graphic depicting the fluorescence distribution of all samples across all experiments for the different panels is shown in **Supplemental Figure 20**. Representative gating strategy and cell hierarchy for each of the panel is shown in **Supplemental Figure 2, S4, S9, S18b and S21b**. Positive gating for each fluorochrome parameter was established using fluorescence minus one (FMO) controls.

### DC phenotyping, data preprocessing and unsupervised clustering analysis

Cryopreserved PBMCs were thawed in 10 batches of 4-5 patients per experiment, with each experiment comprising all visits of interest. 2×10^6^ cells were stained with live/dead FVS780 reagent (BD Biosciences, CA) for 15 min at room temperature, centrifuged and stained with antibodies of DC phenotyping panel (**Supplemental Table 11**) in Brilliant Stain buffer (BD Biosciences, CA) on ice. Subsequently, cells were fixed with 1.5% paraformaldehyde for 10 minutes, washed and acquired on a BD LSRFortessa™ using BD Standardized Application Setup. Samples with viability < 85% were excluded from further analysis. Inter-experiment variation in terms of cell frequency (**Supplemental Figure 21a**) was assessed by running one IC sample in each experiment. Inter-experiment variation for IC resulted to be minimal, while variability among patient samples was significantly larger, as expected for true biological variation (**Supplemental Figure 21a**).

Prior to further analysis, .fcs files were loaded into Flowjo, and HLA-DR+ cells were pre-gated in order to eliminate debris, doublets and dead cells (**Supplemental Figure 21b**). Pre-gated .fcs files were imported into R and logicle transformation was applied using the flowCore package ^48^. Subsequently, a multi-dimensional scaling (MDS) plot was created with R limma package including the expression of all markers, outlier samples with MSD1 < -0.15 were identified and excluded from further analysis (**Supplemental Figure 21c**). Finally, cell populations were identified by unsupervised clustering using FlowSOM ^22^ and ConsensusClusterPlus ^23^ applied to all samples simultaneously. Briefly, a self-organizing map (SOM) was built using the logicle-transformed expression of the lineage markers (Lin, HLADR, CD11c, CD1c, CD141, CD123) with the BuildSOM function, where cells were assigned according to their similarities to 100 grid points of the SOM. Then, metaclustering of the SOM nodes was performed with the ConsensusClusterPlus function. Finally, some metaclusters were manually merged into the cell populations of interest according to their marker expression (**Supplemental Figure 21d, e**).

### Skin Immunofluorescence staining and quantification

6 μm-thick skin sections were fixed with 4% paraformaldehyde (Electron Microscopy Science, PA) and incubated with Image-iT TM FX Signal Enhancer (Invitrogen, CA) for 30 min, to block non-specific antibody binding sites. Next, sections were incubated at 4°C overnight with primary antibodies (Skin immunofluorescent staining panels, **Supplemental Table 11**) in PBS buffer with 5% goat serum. Samples were washed with PBS-Tween 0.1 % and incubated with secondary antibodies (**Supplemental Table 11**) diluted in PBS buffer with 5% goat serum for 1 h at RT. Subsequently, sections were washed with PBS-Tween 0.1% and streptavidin wash added for 30 min at RT. Finally, samples were washed, and nuclear staining was performed by embedding samples in Prolong Gold antifade reagent with DAPI (Invitrogen, CA). Stained sections were evaluated by epifluorescence in a Nikon DS-Qi2 sCMOS Eclipse Ti-2 inverted microscope (Nikon, JP) with a 20× objective running NIS Elements. Image analysis was carried out with NIS Elements software. Briefly, an area of analysis of 750×750 μm was identified and a cellular mask created using bright spot detection with growing enabled in the DAPI image. A minimum intensity threshold was established for CD11c, CD4 or CD8 signals in order to identify the cells of interest. Then, either fluorescence intensity was measured in the cells of interest (CD83 and CD274) or a minimum intensity threshold was established to count the number of double positive cells (IL23, IL17A). A region of interest (ROI) to limit the analysis to the dermis was added for the analysis of dermal DC.

### Statistical analysis

To study the relationship between immune biomarkers and clinical response we modelled the latter first as a continuous variable by dividing PASI at week 12 by PASI at baseline (week 0) and expressing it as a percentage. This variable represents the % residual disease at week 12 e.g a PASI75 R patient has a residual disease of 25%. Correlation between NF-κB nuclear translocation/phosphorylation, frequency of cytokine producing T cell subset or CD274 expression, and residual disease were assessed using univariate linear regression in R for each combination of stimulus, cell type and timepoint.

For significant correlations, differences in NF-κB activation, marker expression or frequencies/counts of immune populations between PASI75 R and NR (binary outcome) were assessed with the non-parametric Mann–Whitney U test or the parametric unpaired t test in R (both two-sided), as appropriate. Normality was assessed with the D’Agostino-Pearson test for groups with more than 8 values; if the group contained fewer than 8 values a non-normal distribution was assumed. The Benjamini–Hochberg approach (False Discovery Rate, FDR) was used to control for multiple tests, as appropriate. For comparisons between two time points involving the same patient, differences were assessed with the non-parametric Wilcoxon test or the parametric paired t test, as appropriate. For comparisons between multiple time points involving the same patient, differences were assessed with the Kruskal–Wallis with Dunn’s multiple comparisons post-test. Receiver operating characteristic (ROC) curve analysis and area under the ROC curve (AUC) comparison were performed using the pROC R package ^49^. For the long-term response analysis, patients that had switched treatment due to inefficacy were considered non-responders. Predictive odds ratio was calculated using the sensitivity and specificity of the assay as reported in ^50^.

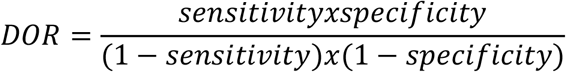

## Supporting information

Sypplemental Material

## Data Availability

All data are included in the manuscript

## Author contributions

RAE performed experiments, analysed and interpreted data and drafted the manuscript. HA and KG processed samples and performed experiments. IT and ZC processed and managed samples. SS performed experiments. CA, JS, ND and EDR analised data and provided statistical advice. HS provided nursing support. AC provided clinical samples. FON conceived and designed the PSORT program and acquired funding. MRB acquired funding and contributed to the design and implementation of the PSORT program. CEMG, JNB, NJR, RBW and CHS conceived and designed the PSORT program, acquired funding, supervised the clinical study and sample collection, and provided insight into data interpretation. PDM interpreted data, designed and supervised the study, and wrote the manuscript. All the authors reviewed the data and contributed to the manuscript.

## Acknowledgments

Supported by the PSORT Consortium, which is in turn funded by a Medical Research Council (MRC) Stratified Medicine award (MR/L011808/1). Partners of the PSORT consortium are AbbVie, the British Association of Dermatologists, Becton Dickinson and Company, Celgene Limited, GlaxoSmithKline, Guy’s and St Thomas’ NHS Foundation Trust, Eli Lilly, Janssen Research & Development, King’s College London, LEO Pharma, MedImmune, Novartis Pharmaceuticals UK, Pfizer Italy, the Psoriasis Association, Qiagen Manchester, Queen Mary University of London, the Royal College of Physicians, Sanquin Blood Supply Foundation, the University of Liverpool, the University of Manchester, and Newcastle University. We particularly acknowledge generous in kind support from the PSORT industrial partner Becton Dickson. All decisions concerning analysis, interpretation, and publication are made independently of any industrial contribution. This research was also supported by the National Institute for Health Research (NIHR) Biomedical Research Centre based at Guy’s and St Thomas’ NHS Foundation Trust and King’s College London, the Newcastle NIHR Biomedical Research Centre, and the NIHR Manchester Biomedical Research Centre. The views expressed are those of the author(s) and not necessarily those of the NHS, the NIHR, or the Department of Health and Social Care. The GTEx Project was supported by the Common Fund of the Office of the Director of the National Institutes of Health, and by NCI, NHGRI, NHLBI, NIDA, NIMH, and NINDS. ND is supported by Health Data Research UK (MR/S003126/1). NJR is supported by the Newcastle NIHR Biomedical Research Centre and the Newcastle NIHR Medtech and In vitro diagnostics Co-operative. CEMG and NJR are NIHR Senior Investigators.

We are grateful to psoriasis patients and healthy volunteers for their participation. We acknowledge the enthusiastic collaboration of the dermatologists and specialist nurses in the UK who recruited to this study. We acknowledge the enthusiastic collaboration of the dermatologists and specialist nurses in the UK who recruited to this study, in particular Prof. David Burden (Western Infirmary, Glasgow), Dr Evmorfia Ladoyanni (Russells Hall Hospital, The Dudley Group NHS Foundation Trust, Dudley), Dr Richard Parslew (Royal Liverpool& Broadgreen University Hospital NHS Trust), and Dr Gayathri Perera (West Middlesex University Hospital, Chelsea and Westminster Hospital NHS Foundation Trust). We thank Alice Russel, Michael Duckworth, Tejus Dasandi, Nadya Dinev, Freya Meynell (London), Tom Ewen, and Dhanisha Lukka (Newcastle) for sample and data management, and Federica Villanova (London) for her contribution to obtain ethical approval. We thank Esme Nichols (Newcastle) for skin sectioning, Susanne Heck, Anna Rose and PJ Chana at the BRC Flow Cytometry Platform at NIHR Guy’s and St Thomas’ Biomedical Research Centre and Virginia Silio and Isma Ali at the Nikon Imaging Centre at Kings College London for technical assistance. We thank Ruth Williams at Novartis for help with clinical data of patients recruited into the Signature study. We thank Dr Brigitta Stockinger for critical reading of the manuscript.

