## Supplementary material for "Enhanced NF-κB signaling in type-2 dendritic cells at baseline predicts non-response to adalimumab in psoriasis": Sypplemental Material

### Supplemental Methods

#### Healthy subjects

Twenty 20 healthy volunteers (mostly Caucasian, 14 males/6 females, age range 27–65 years, mean 44 years, **Supplemental Table 10**) were recruited at Saint John's Institute of Dermatology as age, ethnicity and gender-matched controls to the PSORT combined cohort. Subjects were nominally healthy at the time of sampling.

#### Covariates analysis

The potential contribution of clinical variables (age, gender, ethnicity, smoking, weight, psoriatic arthritis, being biologic naïve, the baseline PASI or the presence of the HLA-C*06:02 allele) to the association between NF-κB phosphorylation and residual disease was assessed by including each variable in turn as a covariate in the linear model. In each case the association remained statistically significant.

#### Genetic analysis

Genetic analysis utilized an expanded version of the PSORT genetic dataset described previously (1) where additional samples were subject to identical eligibility criteria and all samples were of European genetic ancestry. Based on the time windows described in (1), a total of 548 and 763 patients on adalimumab had treatment response measurements at 3m and 6m, respectively.

Genetic association with adalimumab response at the *CD274* gene was assessed in the PSORT genetic dataset as follows. Genotyping array data (1) were imputed to the Haplotype Reference Consortium imputation panel (2) using the Michigan Imputation Server (3). For 355 genetic variants having imputation $R^2 > 0.7$ and minor allele frequency >0.001 in the region 9:5,445,990-5,516,182 (build 37), association with PASI75 and PASI90 response at 3m and 6m were assessed using logistic regression with five ancestry principal components and baseline PASI as covariates. Whole blood eQTL summary statistics from the same region were downloaded from the GTEx Portal on 13 July 2020 (4). A Bayesian test for colocalisation between adalimumab response and whole blood eQTL associations was performed on the set of 59 variants present in both studies using the R package coloc (5) with a prior probability of colocalisation of $10^{-5}$.

### Supplemental Figures

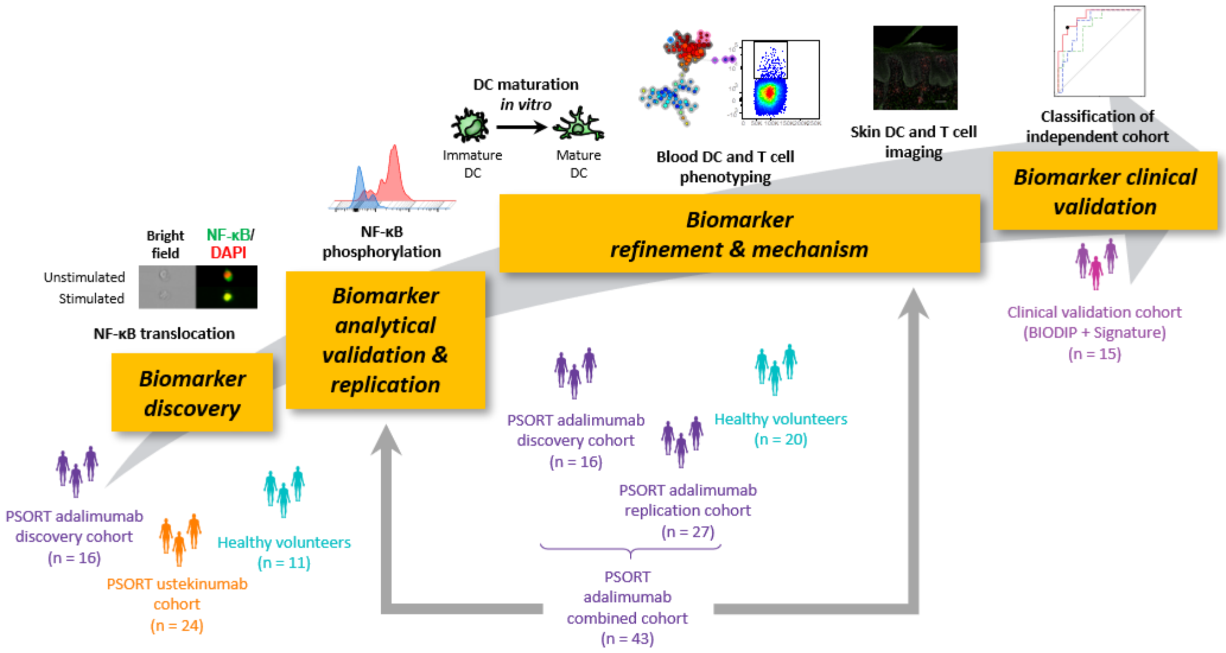


**Supplemental Figure 1. Experimental plan and cohorts used in the study.**

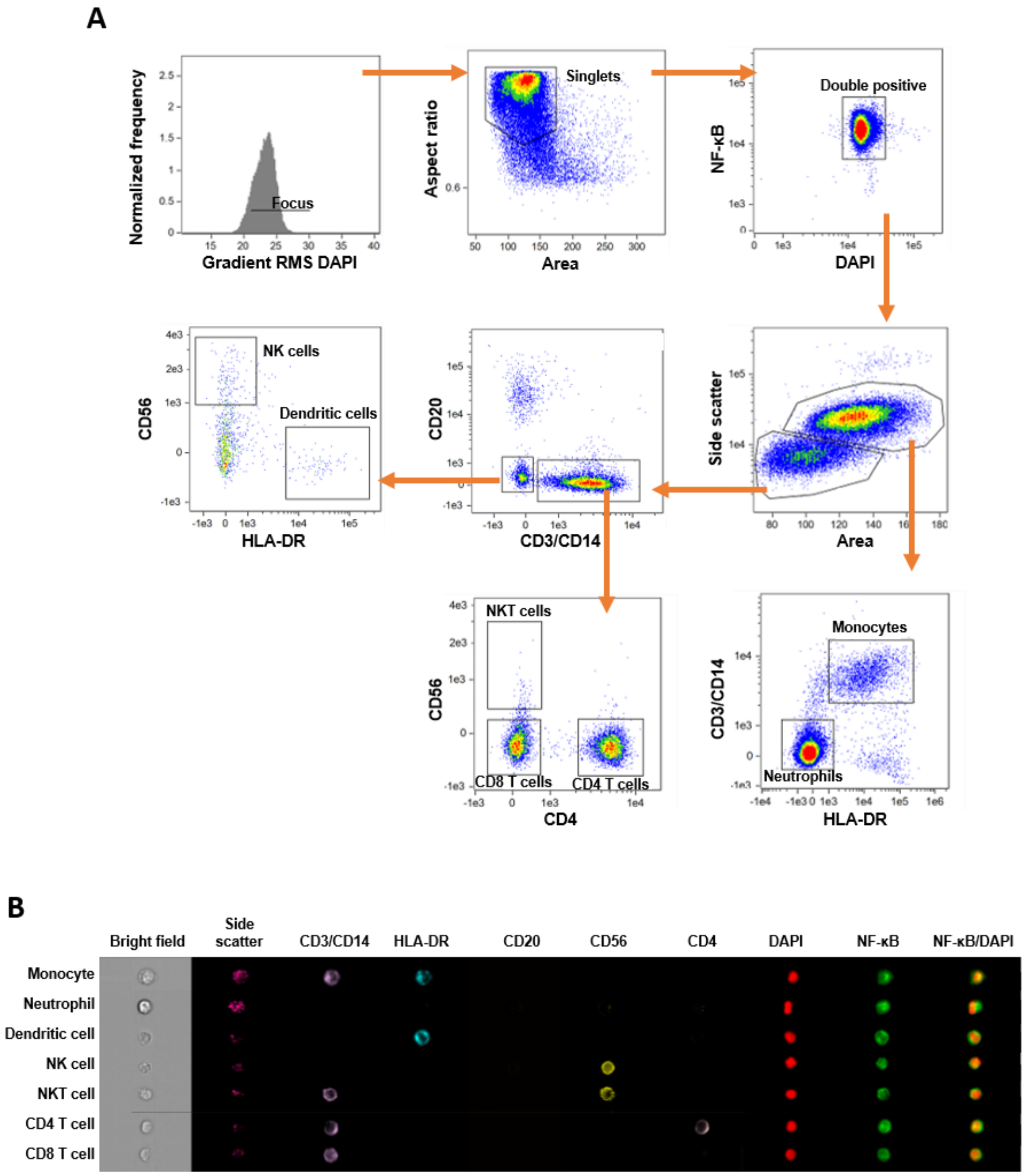


**Fig. S2. Imaging flow cytometry panel. (A)** Representative gating strategy of functional immunophenotyping using imaging flow cytometry. 50.000 focused singlets were acquired. **(B)** Representative image of surface and intracellular markers expression in all cell populations identified (X60 magnification).

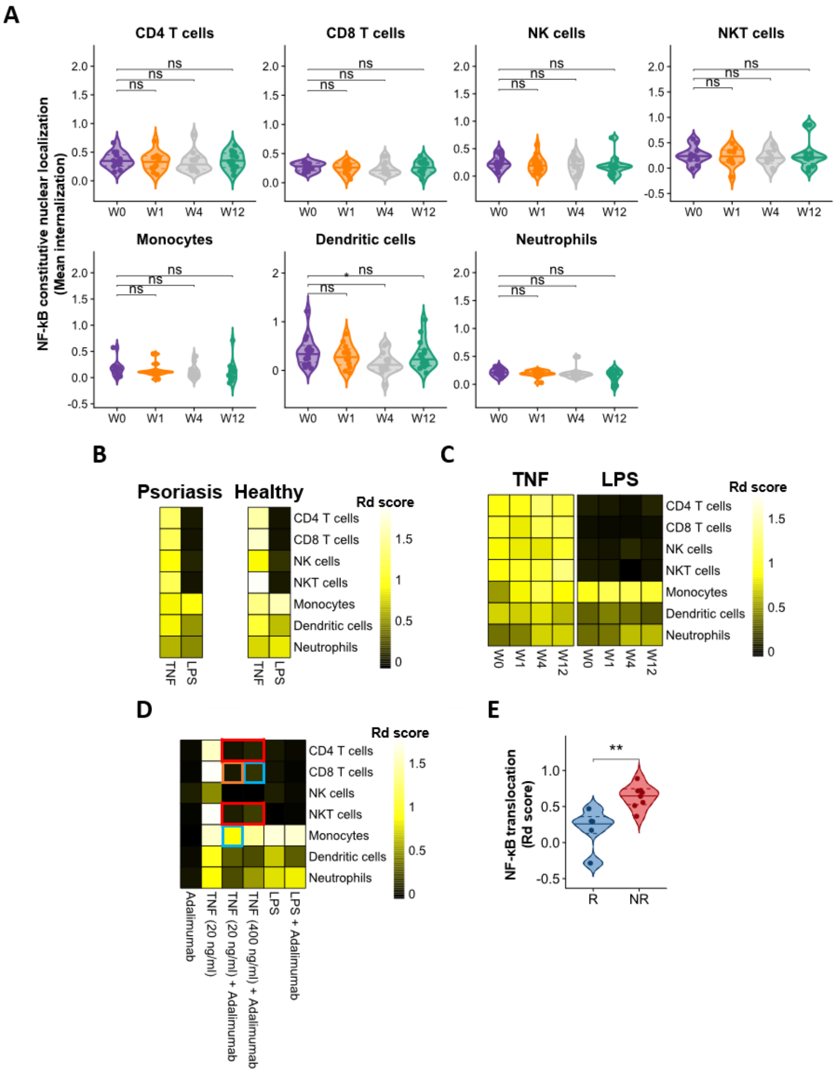


**Supplemental Figure 3. NF-κB nuclear translocation in immune cell subsets.** NF-κB nuclear translocation investigated in whole blood of psoriasis patients before commencing biologic therapy, i.e. at week (w) 0, and at w1, w4 and w12, and of healthy volunteers using imaging flow cytometry. Blood was left unstimulated for constitutive nuclear localization or stimulated with TNF and LPS to induce translocation. **(A)** Violin plot graphs showing constitutive NF-κB nuclear localization (median internalization score) in major immune cell subsets. * $p < 0.05$, Kruskal–Wallis with Dunn's multiple comparisons post-test. **(B)** Heat map showing median TNF and LPS-induced NF-κB translocation in immune cell subsets of psoriasis patients (n=16) and healthy volunteers (n=11). Colour scale indicates the median difference in nuclear translocation of the stimulated condition compared with the unstimulated control (Rd score). **(C)** Heat map showing effect of ustekinumab therapy on NF-κB nuclear translocation in immune cells of psoriatic patients (n=24) **(D)** Heat map showing the effect of in vitro adalimumab to NF-kB nuclear translocation. Whole blood obtained from healthy volunteers was stimulated with either TNF or LPS or left as unstimulated control for 30 min at 37 °C in presence or absence of adalimumab (10 ug/ml). unpaired t test followed by FDE. Red frame *FDR* < 0.0001, orange frame *FDR* < 0.001, blue frame *FDR* < 0.01, n=3. **(E)** Violin plot graph showing LPS-induced NF-kB nuclear translocation (Rd score) in w0 dendritic cells (DC) of PASI90 adalimumab responders (R, blue) and non-responders (NR, red). ** $p < 0.01$, Mann-Witney-U test.

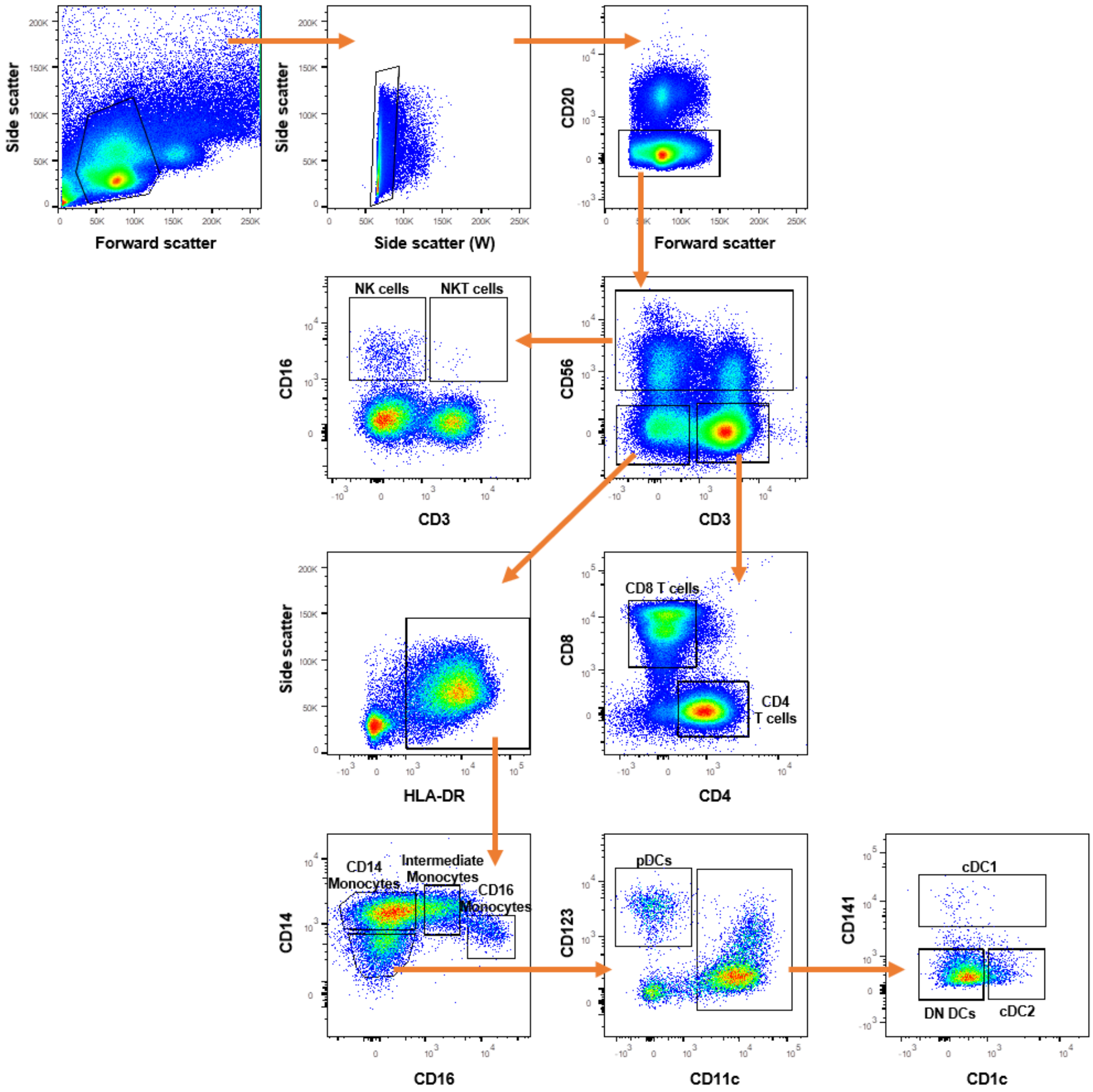


**Supplemental Figure 4. Phospho flow cytometry panel.** Representative gating strategy of phospho flow cytometry panel. 500.000 singlets were acquired

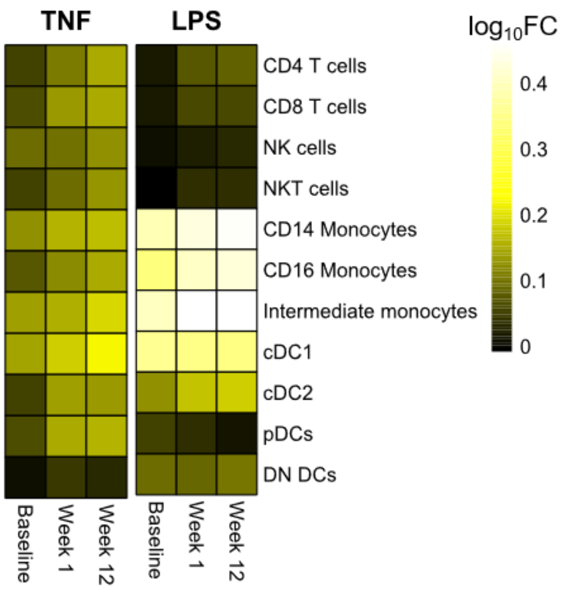


**Supplemental Figure 5. Effect of adalimumab therapy on p65 NF-kB phosphorylation.** PBMC from psoriasis patients at week (w) 0, 1 and 12 of adalimumab therapy were stimulated with either TNF or LPS and NF-κB p65 Ser529 phosphorylation was measured using phospho flow cytometry. Heatmap shows median NF-κB nuclear translocation in immune cell subsets. Colour scale indicates the difference in $log_{10}$ mean intensity of the stimulated condition compared with the unstimulated control. n=43

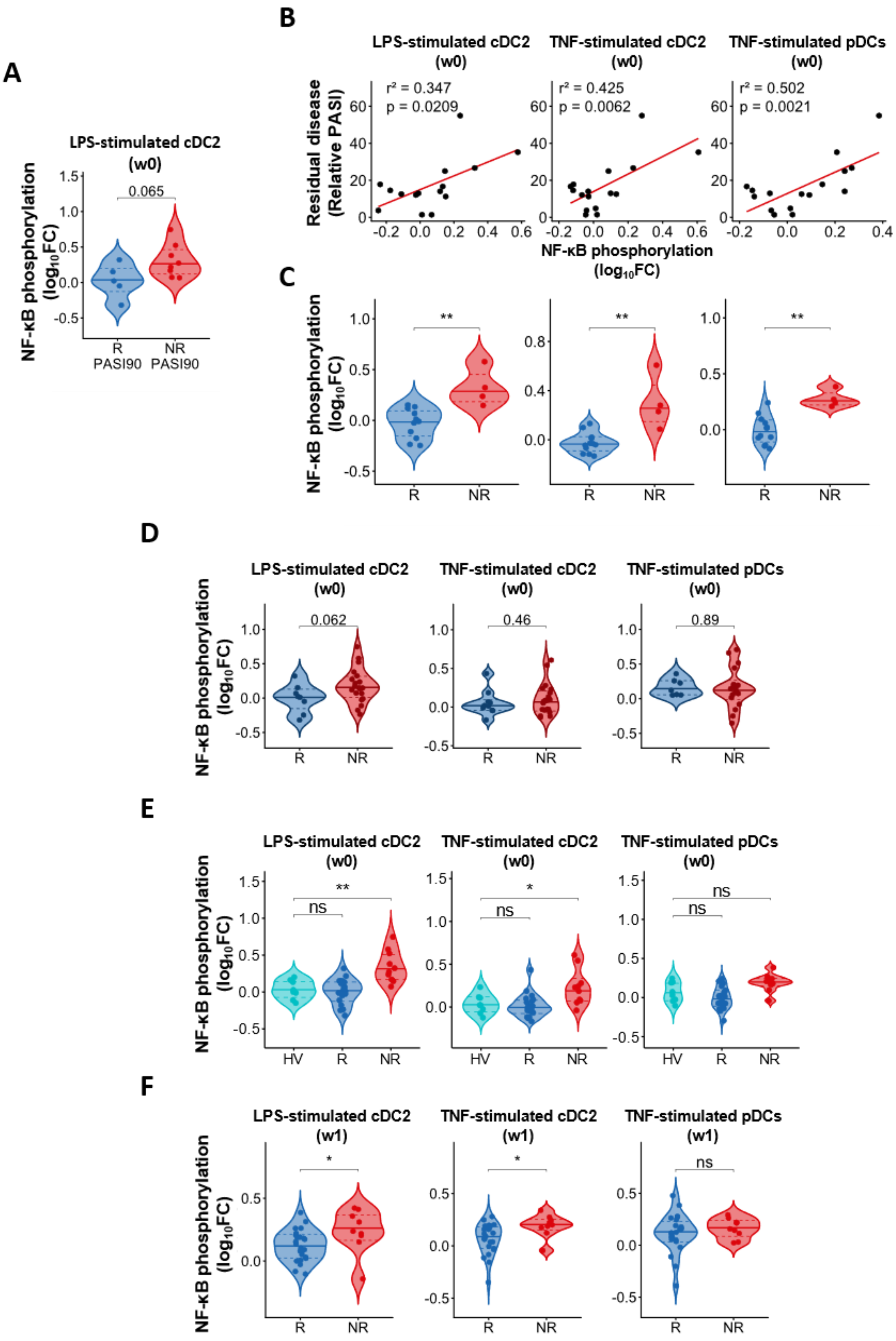


**Supplemental Figure 6. NF-κB phosphorylation in patients undergoing adalimumab therapy.** PBMC from psoriasis patients undergoing adalimumab therapy or healthy volunteers (HV) were stimulated with either TNF or LPS and NF-κB p65 Ser529 phosphorylation was measured using phospho flow cytometry. **(A)** Violin plot graph showing LPS-induced NF-κB phosphorylation in conventional type2 dendritic cell (cDC2) at w0 in PASI90 adalimumab responders (R, blue) and non-responders (NR, red) in discovery cohort (n=13) used for analytical validation. **(B)** Correlation analysis between NF-κB phosphorylation ($log_{10}$FC) at w0 and clinical response expressed as residual disease at w12 (measured as relative PASI, i.e. PASI at w12/ PASI at w0) in DC subsets in PSORT adalimumab replication cohort (n=16) **(C)** Violin plot graphs showing NF-κB phosphorylation at w0 in PASI75 adalimumab R (red) and NR (blue) in DC subsets. **(D)** NF-κB phosphorylation in PASI90 adalimumab R and NR in DC subsets in PSORT adalimumab combined cohort (n=43). **(E)** Violin plot graphs showing NF-κB phosphorylation in DC subsets in HV (cyan, n=20) and at w0 of PASI75 adalimumab R (blue) and NR (red) in PSORT adalimumab combined cohort. **(F)** Violin plot graphs showing NF-kB phosphorylation at w1 in PASI75 adalimumab R (red) and NR (blue) in DC subsets **** $p < 0.0001$, ** $p < 0.01$, * $p < 0.05$, Mann-Whitney U test (A,C,D middle panel, F left and middle panels), unpaired t test (D all but middle panel, F right panel) or One-way ANOVA with Dunnett's multiple comparisons test (E).

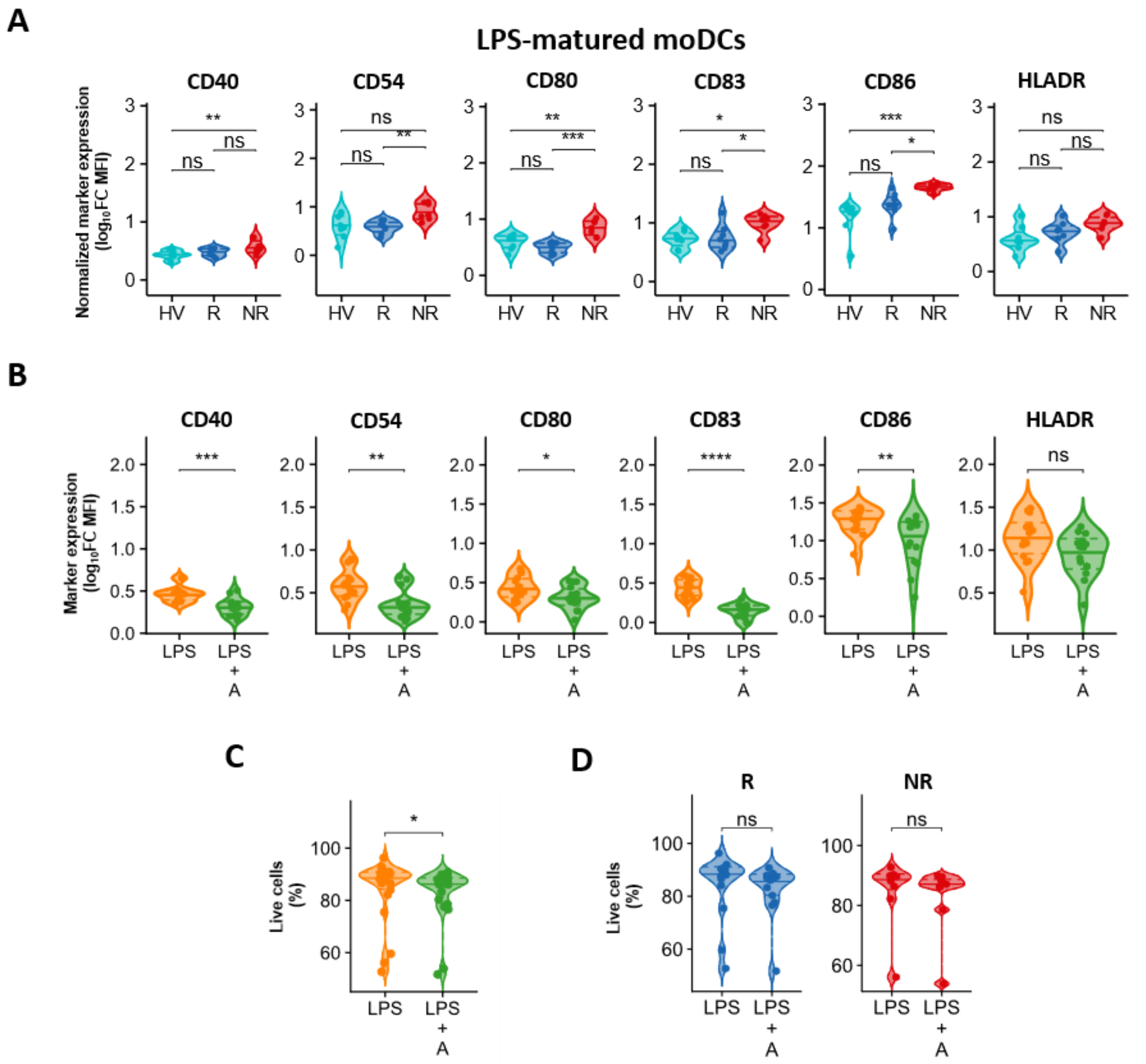


**Supplemental Figure 7. Expression of maturation markers in moDCs.** Monocyte-derived dendritic cells (moDCs) were generated *in vitro* for 6 days from monocytes obtained from healthy volunteers (HV) or psoriasis patients before undergoing adalimumab therapy, and matured with LPS, with and without adalimumab for 2 days. Cells were analysed at day 8 of culture by flow cytometry. **(A)** Violin plot graphs showing expression of LPS-induced co-stimulatory molecule and maturation marker (calculated as $Log_{10}$ Fold Change (FC) Median Fluorescence Intensity (MFI) mature/immature cells) in HV (n=8) and adalimumab PASI75 responders (R, blue, n=8) and non-responders (NR, red, n=7). **(B)** Violin plot graphs showing expression of maturation markers in psoriasis moDCs maturated with LPS alone (LPS) or LPS + Adalimumab (LPS + A) in psoriatic cells. **(C)** Violin plot graph showing psoriasis moDC survival in presence and absence of adalimumab. **(D)** Violin plot graph showing psoriasis moDC survival in presence and absence of adalimumab in PASI75 R and NR. *** $p < 0.001$, ** $p < 0.01$, * $p < 0.05$, Kruskal–Wallis with Dunn's multiple comparisons post-test (A), unpaired t test (B CD40-CD83-HLADR) or Mann-Whitney test (B CD54-CD80-CD86, C, D).

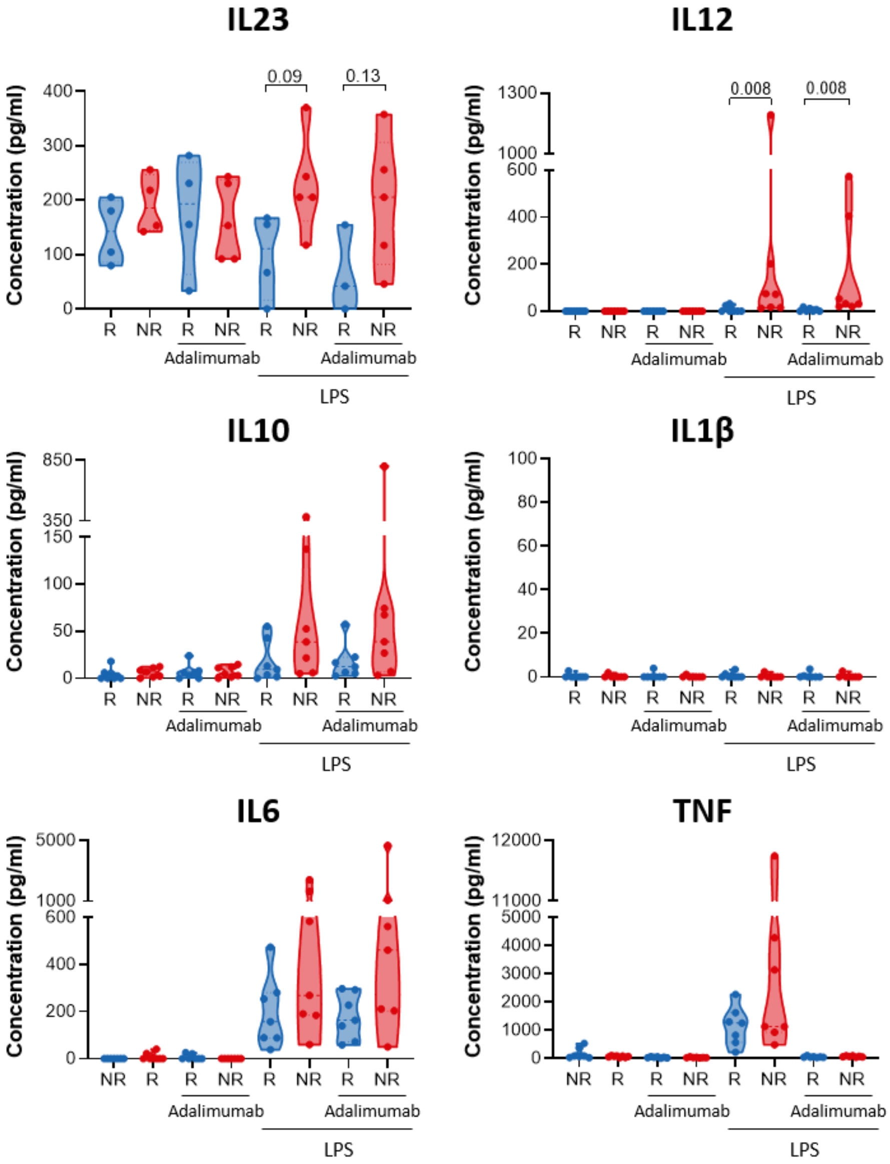


**Supplemental Figure 8. Cytokine production in LPS-matured moDCs.** Monocyte-derived dendritic cells (moDCs) were generated *in vitro* for 6 days from monocytes obtained from psoriasis patients before undergoing adalimumab therapy, and matured with LPS, with and without adalimumab for 2 days. On day 8 of culture supernatants were collected and cytokine levels were measured by Multicytokine bead assay. Kruskal–Wallis with Dunn's multiple comparisons post-test.

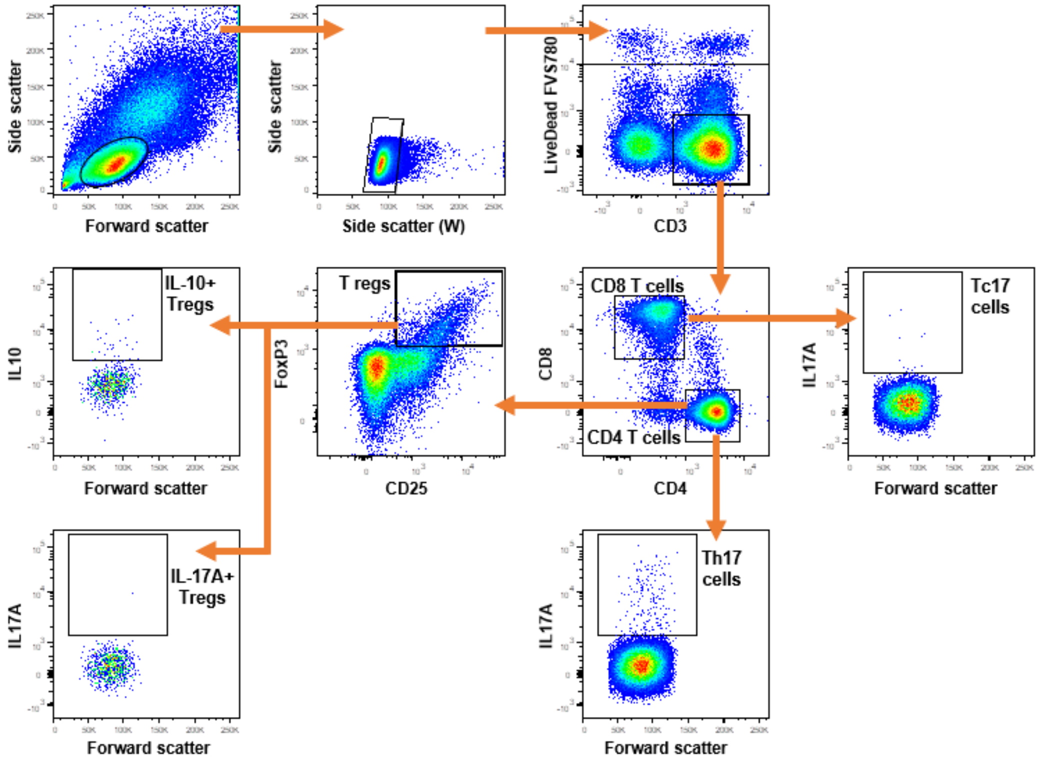


**Supplemental Figure 9. Intracellular cytokine T cell phenotyping panel.** Representative gating strategy of intracellular cytokine T cell phenotyping panel. 250.000 live CD3+ cells were acquired.

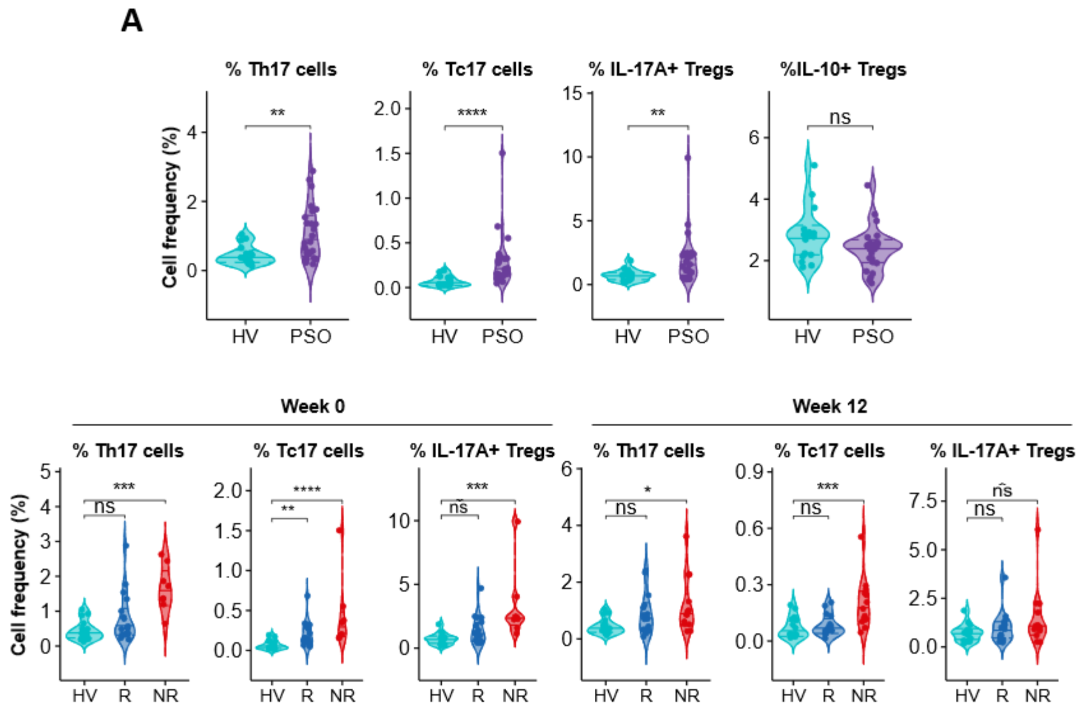


**Supplemental Figure 10. IL17A production in T cells.** PBMCs from healthy volunteers (HV) or psoriasis patient collected before (week 0) and at week 12 of adalimumab therapy were stimulated with PMA/ ionomycin or left unstimulated for 3 hours. Cytokine production was measured by intracellular flow cytometry. **(A)** Violin plot graphs of frequency of IL17A producing cells in CD4+ T (Th17), CD8+ (Tc17) cells and in CD4+CD25+FOXP3+ (Tregs) and of IL-10 producing cells in Tregs in healthy volunteers (cyan, n = 20) and psoriasis patients at w0 (purple, n = 43). **(B)** Violin plot graphs of Th17, Tc17 and IL17+Tregs cells in HV and adalimumab PASI75 responders (R, blue, n=15-18) and non-responders (NR, red, n=8-13) at week 0 and 12 of therapy. **** $p < 0.0001$, *** $p < 0.001$, ** $p < 0.01$, * $p < 0.05$, Mann-Whitey U test (A) or Kruskal–Wallis with Dunn's multiple comparisons post-test (B).

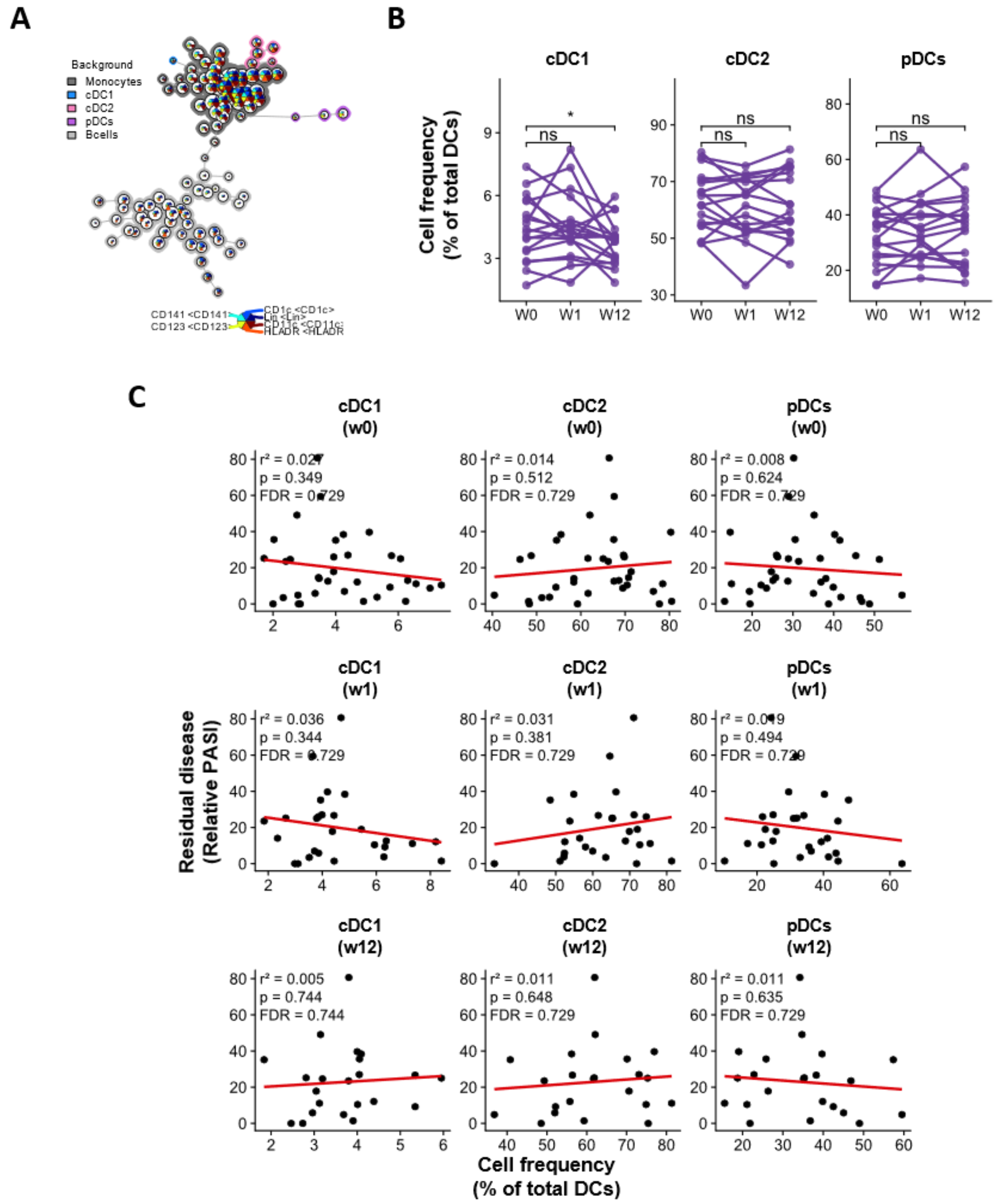


**Supplemental Figure 11. Frequency of dendritic cell subsets does not correlate with clinical response to adalimumab.** Blood dendritic cell (DC) subsets were phenotyped in adalimumab patients before (W0) and after 1 (W1) or 12 weeks (W12) of therapy using flow cytometry. **(A)** DC subsets were identified using unsupervised clustering with FlowSOM and ConsensusClusterPlus. **(B)** Frequency of cDC1, cDC2 and pDCs within total DC at different time points, each line represents one patient (n = 18). **(C)** Correlation analysis between the frequency of DC subsets within total DCs at different time points and residual disease at week 12 using a generalized linear model (*n* = 43). * $p < 0.05$, paired t test.

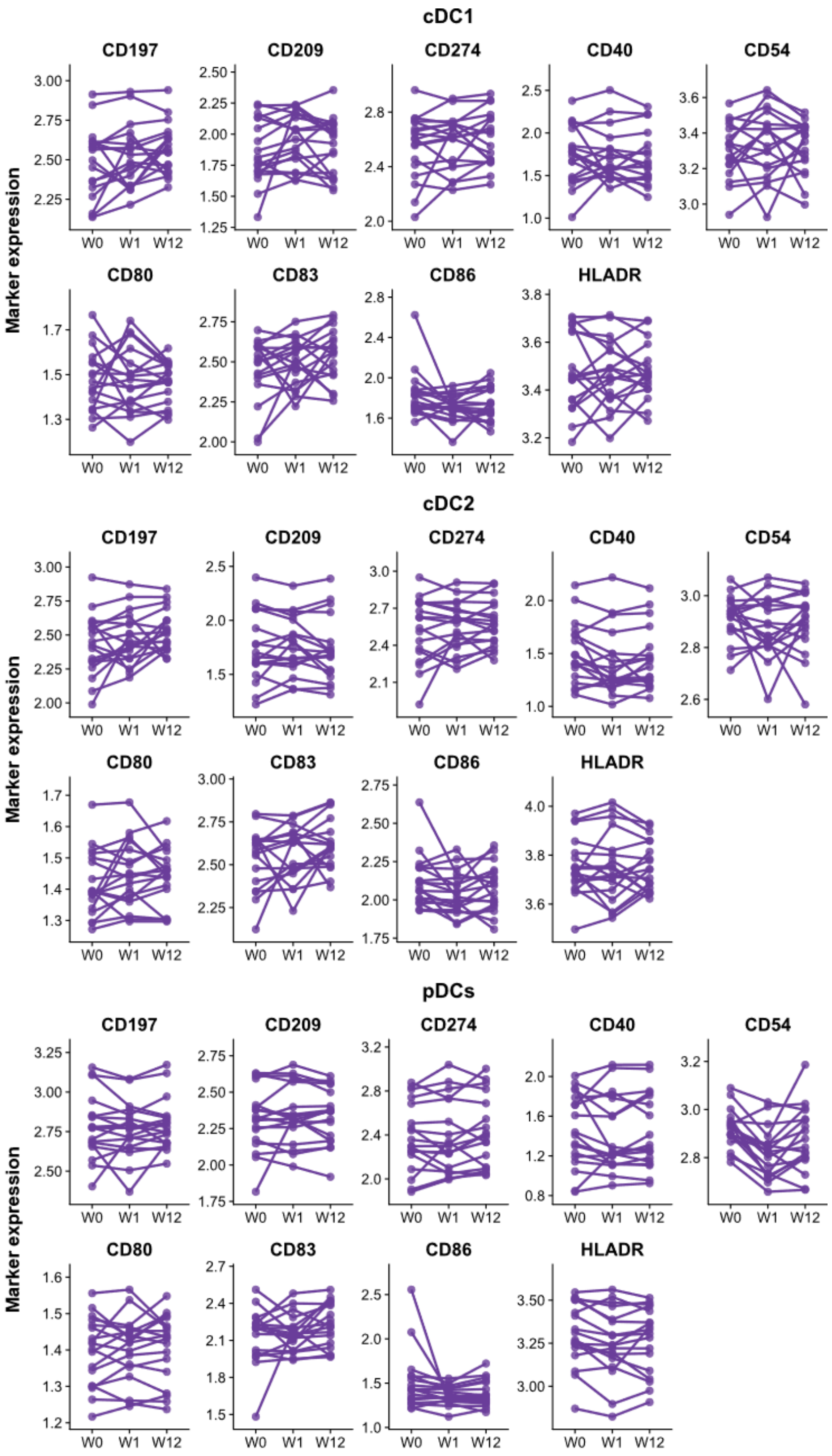


**Supplemental Figure 12. Phenotype of dendritic cell subsets during adalimumab therapy.** Blood DC subsets were phenotyped in adalimumab patients before (W0) and after 1 (W1) or 12 weeks (W12) of therapy using multi-parameter flow cytometry and analysed by unsupervised clustering with FlowSOM and ConsensusClusterPlus. Expression of surface markers of maturation, activation in cDC1, cDC2 and pDCs at different time points, each line represents one patient (*n* = 18). No comparison is FDR significant.

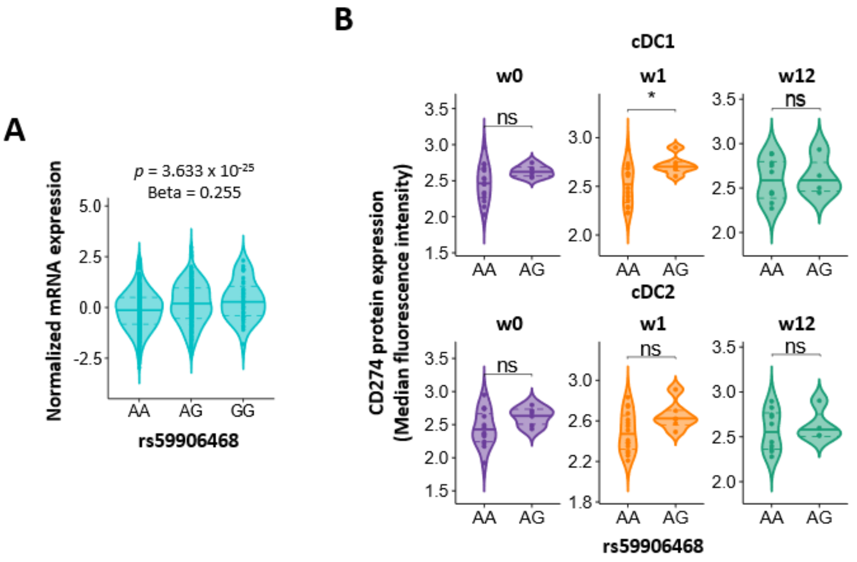


**C**

| Outcome | SNP | | Effect allele | OR | L95 | U95 | Pval |
|---|---|---|---|---|---|---|---|
| PASI75 at 3m | rs72705444 | *Top response SNP* | C | 0.55 | 0.38 | 0.80 | 1.6E-03 |
| | rs59906468 | *Top coloc SNP* | G | 1.34 | 0.96 | 1.86 | 0.08 |
| PASI75 at 6m | rs2297136 | *Top response SNP* | G | 0.64 | 0.51 | 0.81 | 1.5E-04 |
| | rs59906468 | *Top coloc SNP* | G | 1.56 | 1.15 | 2.12 | 4.1E-03 |
| PASI90 at 3m | rs7023227 | *Top response SNP* | T | 0.72 | 0.56 | 0.92 | 0.010 |
| | rs59906468 | *Top coloc SNP* | G | 1.22 | 0.91 | 1.65 | 0.187 |
| PASI90 at 6m | rs2297136 | *Top response SNP* | G | 0.63 | 0.51 | 0.77 | 8.8E-06 |
| | rs59906468 | *Top coloc SNP* | G | 1.52 | 1.18 | 1.96 | 1.2E-03 |

**D**

| | | Colocalisation posterior probabilities | | | | | Likely shared variant | |
|---|---|---|---|---|---|---|---|---|
| **Outcome** | **Number of SNPs in region modelled** | **Model 0: no causal variant** | **Model 1: causal variant for eQTL only** | **Model 2: causal variant for response only** | **Model 3: distinct causal variants** | **Model 4: shared causal variant** | **SNP** | **Posterior probability** |
| PASI75 at 3m | 59 | 0.00 | 0.86 | 0.00 | 0.01 | 0.14 | rs59906468 | >0.999 |
| PASI75 at 6m | 59 | 0.00 | 0.55 | 0.00 | 0.01 | 0.44 | rs59906468 | >0.999 |
| PASI90 at 3m | 59 | 0.00 | 0.90 | 0.00 | 0.01 | 0.09 | rs59906468 | >0.999 |
| PASI90 at 6m | 59 | 0.00 | 0.30 | 0.00 | 0.04 | 0.66 | rs59906468 | >0.999 |

**Supplemental Figure 13. Genetic basis for the effect of CD274 on response to adalimumab. (A)** Violin plot for *CD274* mRNA expression in whole blood stratified by rs59906468 genotype in GTEX dataset (n = 669). *p* value and beta effect of the GTEX eQTL are reported. **(B)** Violin plot for CD274 protein expression in adalimumab combined cohort (n=22) stratified by rs59906468 genotype in DC subsets at Week0, 1 and 12. * p <0.05 **(C)** Genetic association analysis of *CD274* region and response to adalimumab assessed as PASI75 or PASI90 at 3 or 6 months. **(D)** Bayesian colocalization analysis of rs59906468 effect on both *CD274* mRNA expression and adalimumab response.

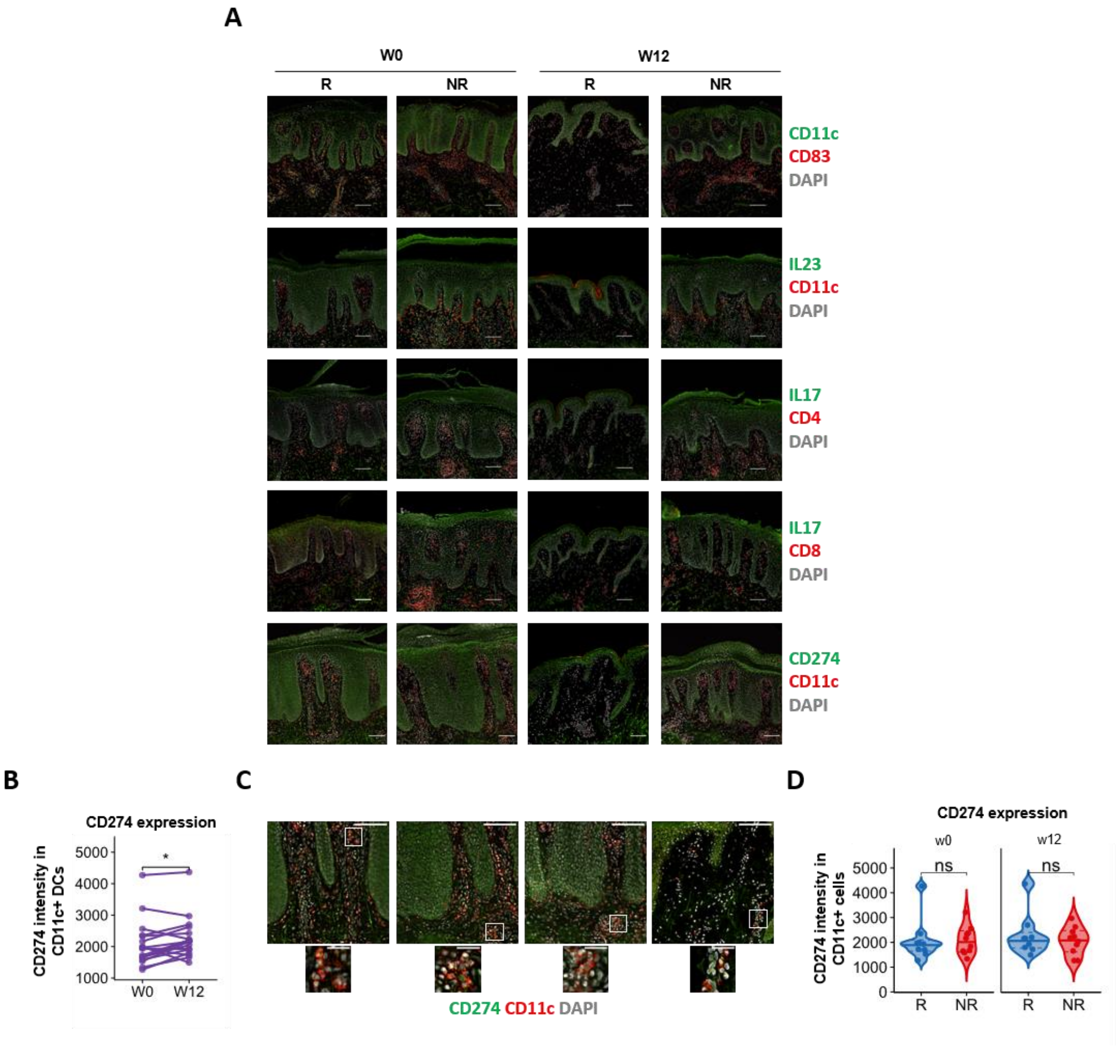


**Supplemental Figure 14. Immunofluorescence analysis of psoriasis skin lesions.** Lesional psoriasis skin was analysed by fluorescence microscopy in PASI75 responder (R) and non-responder (NR) patients before the start of adalimumab (w0) and after 12 weeks (w12) of therapy for CD83 and CD274 expression in CD11c+dermal dendritic cells (dDC), number of IL23+ dendritic cells and number of Th17 and Tc17 cells. **(A)** Representative fields of vision for immunofluorescence analysis. **(B)** Expression of CD274 in CD11c+ dDC at different time points. Each line represents one patient. **(C)** Representative immunofluorescence images of psoriasis skin showing CD274 expression in CD11c+ DC in PASI75 adalimumab R and NR. **(D)** Violin plot graphs of CD274 expression in dermal DC in adalimumab R (blue, n=9-10) and NR (red, n=9-10) at w0 and w12. * $p < 0.05$, paired t test (B), unpaired t test (D). Scale bars in B, E: 100 µm (overview) and 25 µm (insets).

**A**

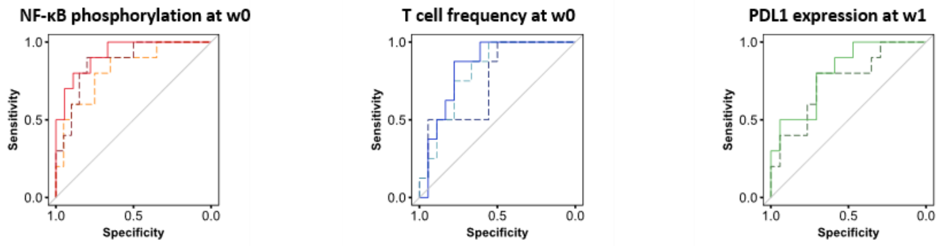


| | AUC | CI | *p* |
|---|---|---|---|
| LPS-cDC2 | 0.922 | 0.826 - 1 | 7.528 x10$^{-5}$ |
| TNF-pDCs | 0.875 | 0.746 – 1 | 4.997 x10$^{-4}$ |
| TNF-cDC2 | 0.825 | 0.665 - 0.984 | 0.003 |

| | AUC | CI | *p* |
|---|---|---|---|
| Th17 | 0.840 | 0.686 – 0.994 | 0.005 |
| IL17A+ Tregs | 0.813 | 0.646 – 0.978 | 0.011 |
| Tc17 | 0.750 | 0.545 – 0.954 | 0.046 |

| | AUC | CI | *p* |
|---|---|---|---|
| cDC2 | 0.806 | 0.638 – 0.973 | 0.008 |
| cDC1 | 0.747 | 0.549 – 0.944 | 0.035 |

**B**

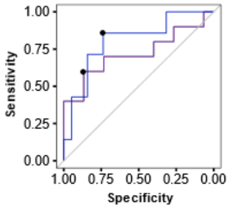


| | AUC | CI | *p* | Sensitivity | Specificity |
|---|---|---|---|---|---|
| 6m | 0.80 | 0.6024 - 1 | 0.018 | 85.7% | 73.7% |
| 12m | 0.72 | 0.4869 – 0.9531 | 0.071 | 60% | 86.7% |
| 24m | 0.72 | 0.4869 – 0.9531 | 0.071 | 60% | 86.7% |

**Supplemental Figure 15. Evaluation of predictive biomarkers of clinical response to adalimumab. (A)** Receiver operator characteristic (ROC) curves of generalized linear models for nine immune traits correlating with response predicting PASI75 outcome at w12 in adalimumab patients (combined cohort). **(B)** ROC curves of LPS-induced NF-κB phosphorylation in cDC2 at w0 predicting PASI75 outcome at 6, 12 and 24 months in adalimumab patients (combined cohort). p values are based on two-tailed Mann–Whitney tests vs an AUC of 0.5.

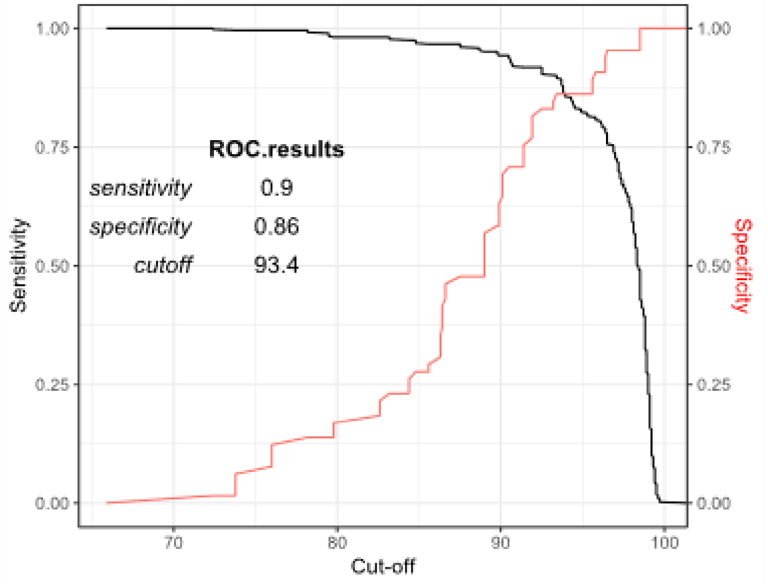


**Supplemental Figure 16. Quality control (QC) of phospho flow cytometry experiments by ROC curve analysis for cell viability cut-off.** Cryopreserved PBMC samples were thawed and rested overnight, viability was measured the following day using a NucleoCounter NC-200 and samples were stimulated with either TNF, LPS or IL-1β for 15 minutes at 37 °C. Phosphorylation of NF-κB p65 at SEr529 was assessed using phospho flow cytometry. Receiver Operating characteristic (ROC) curve identifying viability cut-off which best discriminate good quality samples, set as those with frequency of CD14+ cells >5% and LPS-induced NF-kB phosphorylation in CD14+ cells >0.2$\log_{10}$FC, from low quality samples to be excluded from the analysis.

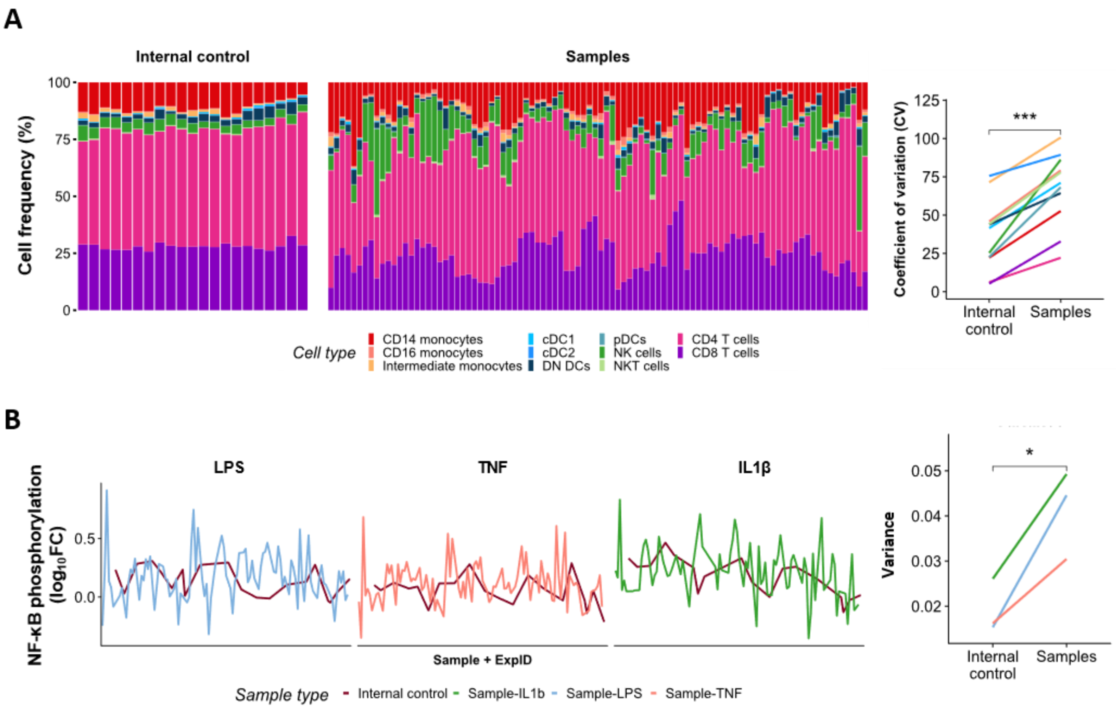


**Supplemental Figure 17. Assessment of phospho flow cytometry inter-experiment variability (A)** Stacked bar plot of cell frequency for populations of interest in the internal control (IC) and test samples (left). Coefficient of variation in IC and test samples, each line denotes a cell type. ** $p < 0.01$. Wilcoxon test. **(B)** NF-κB phosphorylation induced by various stimuli in conventional type 2 dendritic cell (cDC2) in IC and test samples (left). Variance observed in IC and test samples, each line denotes one stimulus. * $p < 0.05$. Paired t test.

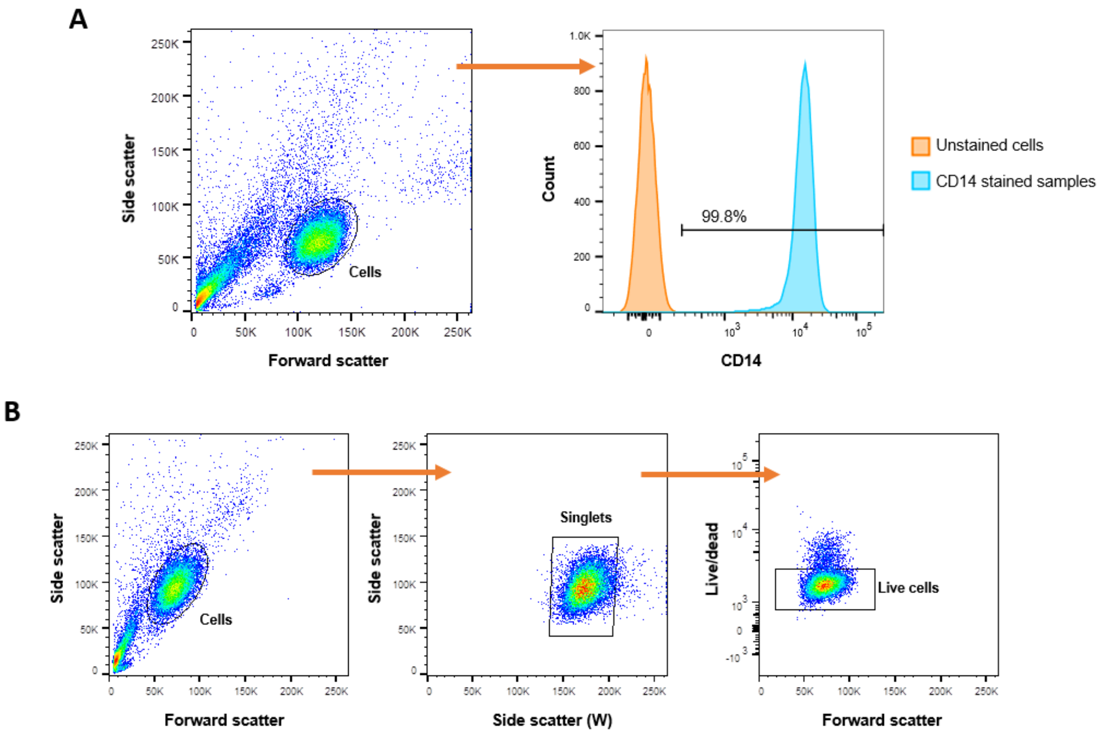


**Supplemental Figure 18. Generation of monocyte-derived dendritic cells (moDC). (A)** Representative gating strategy and histogram showing the purity of isolated CD14+ monocytes **(B)** Representative gating strategy for moDCs experiments.

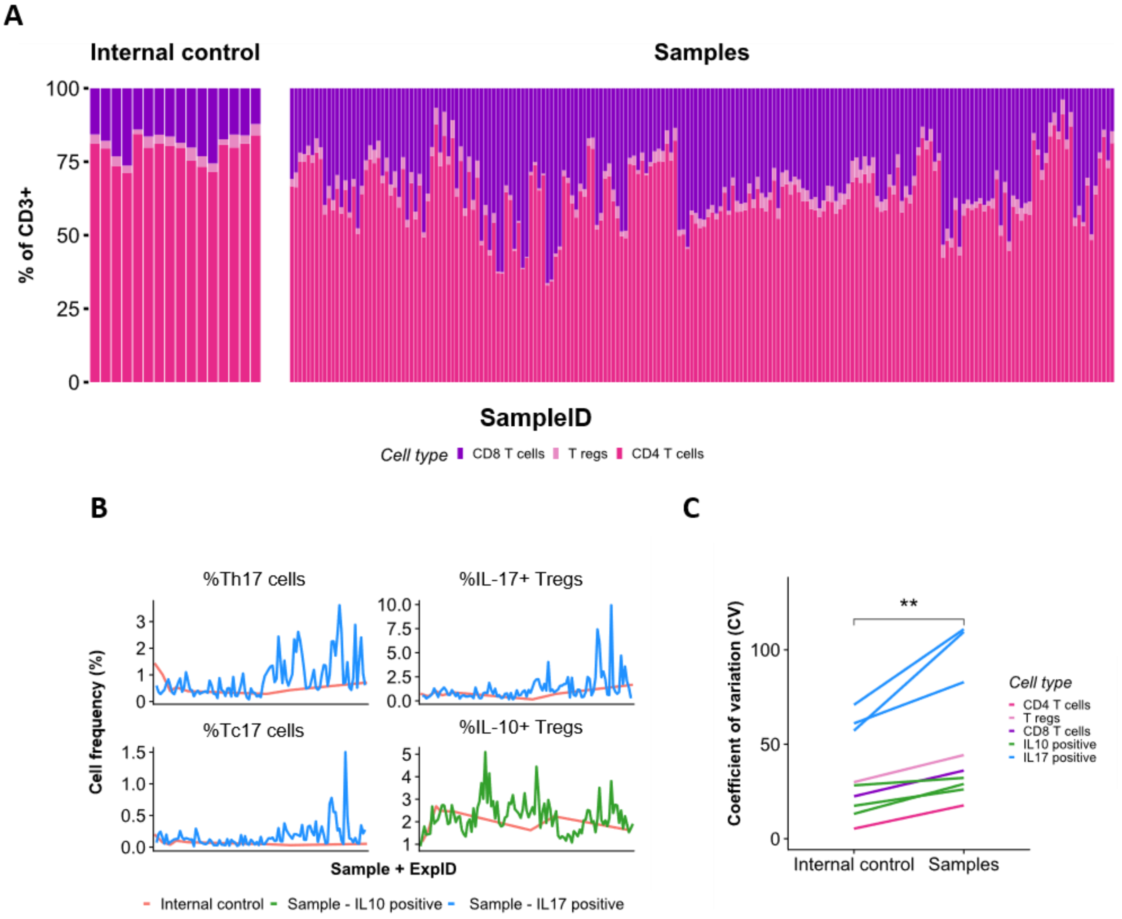


**Supplemental Figure 19. Assessment of T cell phenotyping inter-experiment variability (A)** Stacked bar plot of cell frequency for populations of interest in the internal control (IC) and test samples. **(B)** Frequency of cytokine producing cells in IC and test samples. **(C)** Coefficient of variation of cell frequencies in A and B in IC and test samples, each line denotes a cell type. ** $p < 0.01$. Wilcoxon test.

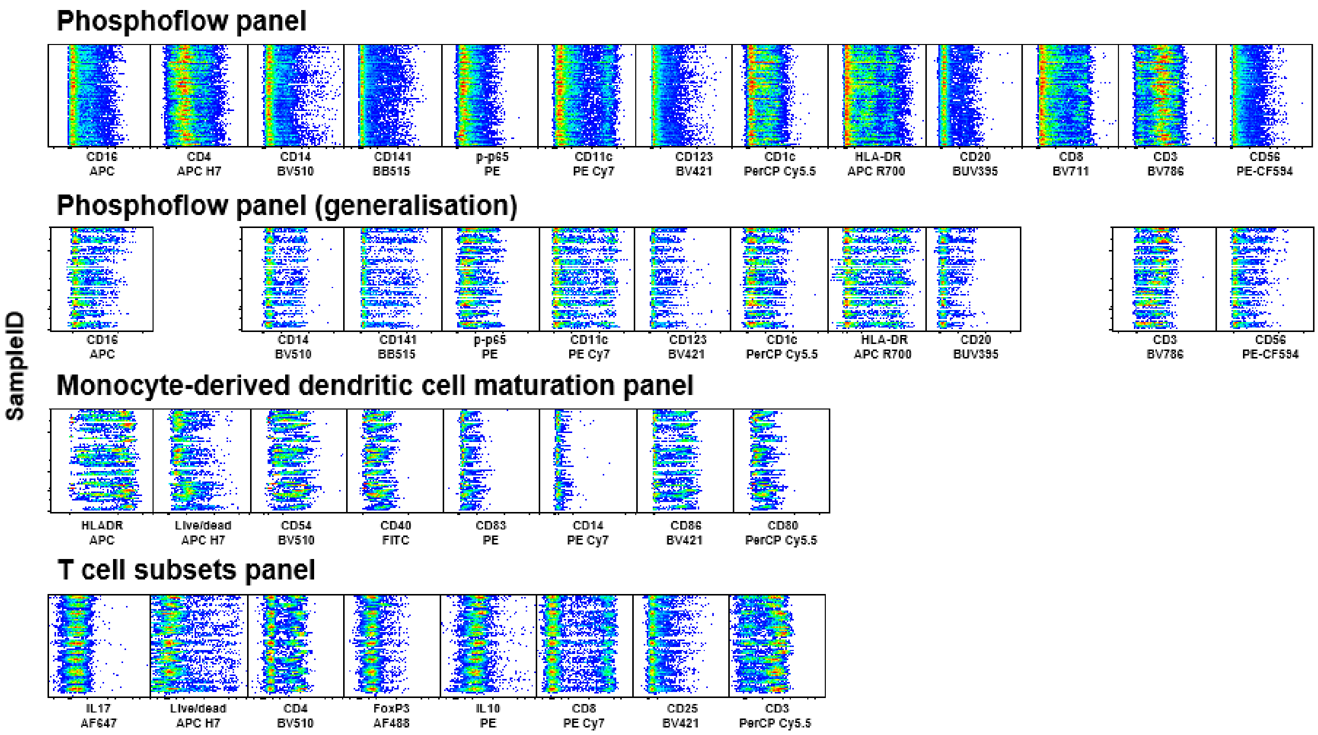


**Supplemental Figure 20. Fluorescence distribution of each flow cytometry panel for all samples**, related to experimental procedures. For each of the four panels, 1000 events from each data file for each of the samples analysed were merged into a single file. Each of the fluorescent markers used in the panel (abscissa) are shown in separate graphics, plotted against all samples (ordinate). Antibody panels were identical throughout the study.

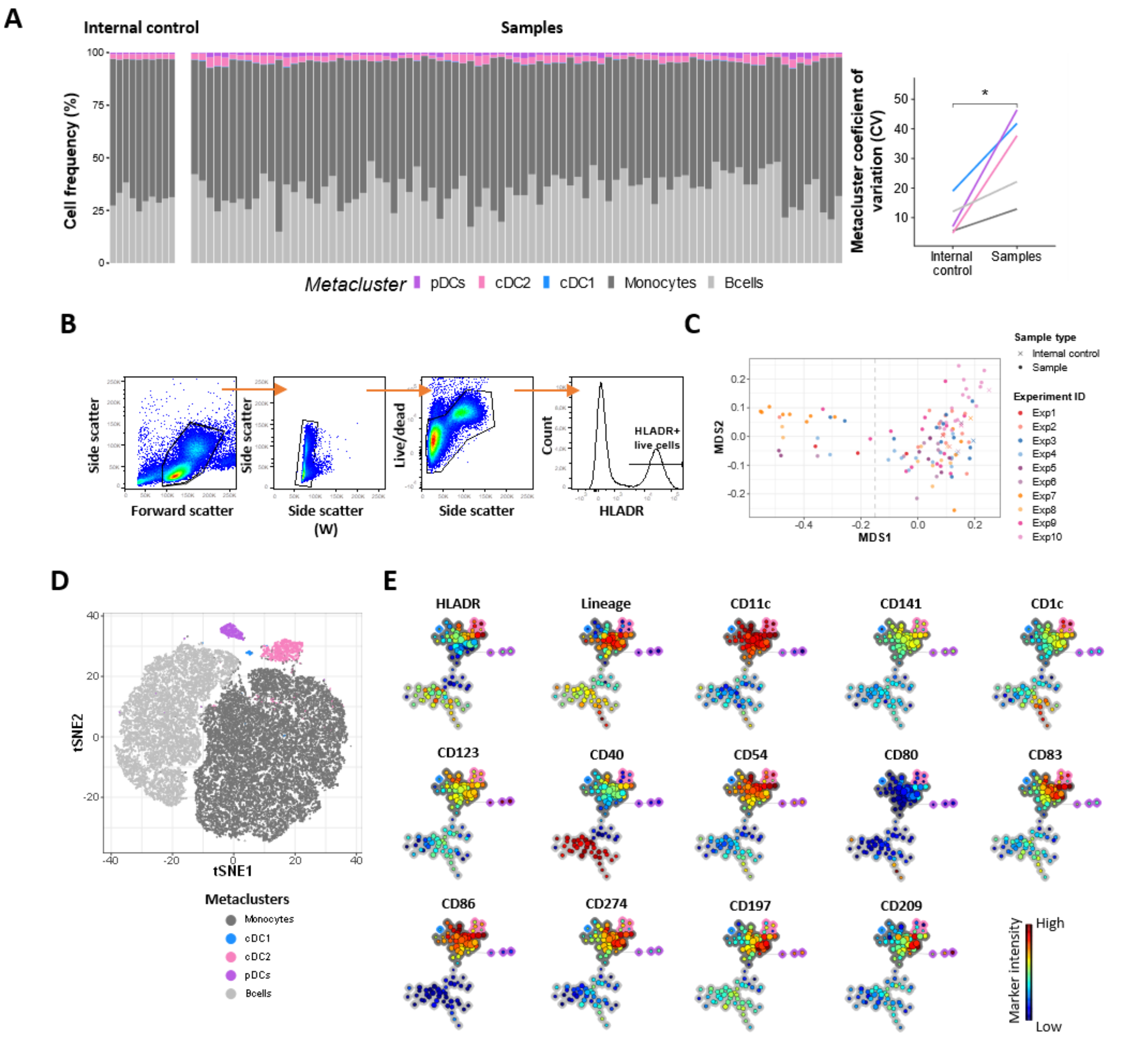


**Supplemental Figure 21. Immunophenotyping and unsupervised clustering of dendritic cell. (A)** Stacked bar plots of frequency of the metaclusters in the internal control (IC) and test samples (left) and coefficient of variation of cell frequencies in A and B in IC and test samples, each line denotes a cell subset (right). **(B)** Manual pre-gating of live HLADR+ cells. **(C)** Multi-dimensional scaling (MDS) plot for outlier identification. **(D)** t-SNE plot based on the logicle-transformed expression of the 6 lineage markers. 2000 cells were randomly selected from each sample. Colours represent the metaclusters. **(E)** Relative marker expression for the SOM nodes. Background colors represent the metaclusters. * $p < 0.01$. Wilcoxon test.

**List of Supplemental tables**

Supplemental Table 1. Effect of adalimumab on NF-κB translocation in immune cells.

Supplemental Table 2. Correlation analysis between NF-κB translocation in immune cells and response to adalimumab.

Supplemental Table 3. Effect of adalimumab on NF-κB phosphorylation in immune cells.

Supplemental Table 4. Correlation analysis between NF-κB phosphorylation in dendritic cells and response to adalimumab.

Supplemental Table 5. Covariate analysis for the significant correlations between NF-κB phosphorylation and response to adalimumab.

Supplemental Table 6. Correlation analysis between T cell subset frequency and response to adalimumab.

Supplemental Table 7. Effect of adalimumab on dendritic cell phenotype.

Supplemental Table 8. Correlation analysis between dendritic cell phenotype and response to adalimumab.

Supplemental Table 9. Differences in dendritic cell phenotype between adalimumab PASI75 responders and non-responders.

Supplemental Table 10. Patient information and demographics.

Supplemental Table 11. Antibodies used in the study.
